# A Network Analysis of Built Environment Features and Depressive Symptoms over an 18-year period

**DOI:** 10.64898/2026.03.18.26348702

**Authors:** Faye Sanders, Lucy Waldren, Vilte Baltramonaityte, Esther Walton

## Abstract

Although the built environment has been identified as a risk factor for depressive symptoms, it is unclear whether these associations are driven by specific environmental features and whether they remain stable over time.

In 10,310 ALSPAC women living in Bristol city, we conducted preregistered network analyses to investigate cross-sectional and longitudinal associations between built environment features (e.g., population density, green space and walkability) and depressive symptoms (at ages 28, 32 and 48 years).

Contrary to our hypotheses, associations between individual built environment variables and depressive symptoms were consistently weak. Exploratory factor analyses indicated a built environment factor associated with depressive symptoms at baseline (β = 0.148, *p* < .001) and 4-year follow-up (β = 0.114, *p* = .011), but not at 18-year follow-up (β = -0.005, *p* = .950).

These findings suggest the combined influence of built environment features may explain depressive outcomes better than individual built environment measures alone.

## Introduction

Urbanicity has been identified as a risk factor for poor mental health, including depression, for some time ^1^. Generally, urbanicity refers to multiple urban conditions of a city at a moment in time, including population density; high quantity of educational facilities, transport, and government services; and low green space ^2^. However, research investigating links between urbanicity and mental health most commonly use population density as a single-measure proxy for urbanicity ^3^. However, using population density as a single proxy for multiple domains of the built environment can limit the specificity and effectiveness of urban planning initiatives targeted at improving public mental health, as it remains unclear whether it is population density driving these associations or other closely associated and more modifiable factors.

Green space is one such closely associated feature which has been paid considerable attention in the literature. For example, green space associates with various positive mental health outcomes ^4,5^. Similarly, access to green space is often included as a component of walkability, a term used to describe how easy and accessibly an area can be navigated as a pedestrian ^6^. Higher levels of walkability have been found to predict lower rates of depression across a wide range of contexts ^7–9^.

Transport infrastructure, facility and building density have also been linked to mental health ^10^, although overall, these associations have been comparatively less well researched. For example, experiencing transport congestion consistently associates with psychophysiological distress and poor mental health ^11,12^. Preliminary evidence suggests greater facility density of medical, fitness, entertainment and transport facilities appear to predict better mental health ^13^. However, whilst there is some evidence to suggest these features associate with mental health, it still remains unclear 1) whether these associations are mainly driven by population density and 2) how stable they are over time.

Regarding the first point, current methodologies of regression analyses have been useful in understanding the extent to which the built environment associates with depression ^11,14–18^. However, relationships between individual built environment features are complex and interrelated ^19^. Unlike regression, which primarily focuses on direct relationships between predictors and an outcome, network analysis reveals how variables interact within a broader system ^20^. A network analysis can also pinpoint specific aspects of the built environment most strongly linked to depression whilst modelling interconnections among built environment variables. As such, network analyses have key advantages in helping to provide a deeper understanding of how multiple built environment features holistically and independently of each other associate with depression.

Regarding the second point (stability over time), our understanding of relationships between the built environment and depression is largely underpinned by cross-sectional research. It remains unclear as to how long these associations remain present, and whether stability varies across built environment features. This understanding is critical for identifying where investments in the built environment may be able to generate long-lasting health impacts.

To address these challenges, we used network analysis to investigate relationships between a broad range of built environment features (including but not restricted to population density) and depressive symptoms both cross-sectionally and longitudinally over an 18-year follow-up period. We aimed to investigate 1) how built environment features relate to each other; 2) which built environment features have the strongest associations with depressive symptoms and 3) how these associations change over time.

We preregistered our study prior to analysis on the Open Science Framework (https://osf.io/mg7z6). With regards to aim 1, we expected population density to have the greatest expected influence across networks, due to its role as a built environment proxy measure in previous literature. We also hypothesised two largely separate clusters would form within built environment nodes in the network: one including transport-related variables (e.g., connectivity density, total traffic load on major roads) and another one comprising facility/density-related variables (e.g., facility density, facility richness, building density).

With regards to aim 2, we expected depressive symptoms to be positively associated with total traffic load on major roads and distance to nearest road, due to preliminary evidence (e.g. ^21,22^) suggesting positive associations between proximity to high-traffic roads and poor health. We further expected depressive symptoms to be negatively associated with green space and walkability, due to previous research identifying positive relationships between physical activity, green space and mental health. We did not hypothesise which built environment variable would have the greatest association (edge) with depression, due to a lack of previous research comparing different measures.

With regards to aim 3, we hypothesised associations (edges) identified in aim 2 would remain present over time but weaken as influencing features change across time. As a result, we also hypothesised that the closeness values of depressive symptoms would weaken over time, whilst the general patterns of associations within the network would remain stable.

## Results

### Descriptive Statistics

Participants generally lived in areas of Bristol city where on average half the area was covered in buildings. In addition, the average green space level (mean NDVI = 0.42, SD =0.09) was comparable to those of San Francisco (mean NDVI =∼0.47), and just below average green space across Climate 40 network cities (mean NDVI = 0.54), including New York City and Tokyo ^23^. The mean population density was 4,857.80 people per square kilometer (SD = 2756.99), which is less than the average population density of Greater London of 5,690.00 people per square kilometer ^24^. All participants were female and aged between 15-44 years at enrolment (**Table 1**).

**Table 1.** Study descriptives for 10,310 women who had completed questionnaire items for their depressive symptoms at enrolment and/or at the ∼4-year or ∼18-year follow-up (imputed data). ^1^ Depressive symptoms measured at enrolment and a 4-year follow-up used the Crown Crisp Experiential Index. Depressive symptoms measured at ∼18-year follow-up used the 36-Item Short Form Survey mental health survey. ^2^ Ethnicity (e.g., white, black/Caribbean, black/African, black/other, Indian, Pakistani, Bangladeshi, Chinese, any other ethnic group) was self-reported at study enrolment. SD = standard deviation; SES = socioeconomic status.

|  | Enrolment | 4- year follow-up | 18-year follow-up |
| --- | --- | --- | --- |
| **Age** |  |  |  |
| Mean (SD) | 28.12 (4.84) | 31.53 (4.74) | 47.26 (4.60) |
| Min, Max | 15.00, 44.00 | 18.67, 53.73 | 34.00, 63.00 |
| **Ethnicity** |  |  |  |
| White | 97.77% |  |  |
| Non-White | 2.23% |  |  |
| **Built environment** |  |  |  |
| *Facility density (no. of facilities per km<sup>2</sup>)* |  |  |  |
| Mean (SD) | 32.99 (42.55) |  |  |
| Min, Max | 0.00, 200.00 |  |  |
| *Facility Richness (no. of different facility types in the 300m buffer available, divided by the maximum potential number of facility types)* |  |  |  |
| Mean (SD) | 0.07 (0.07) |  |  |
| Min, Max | 0.00, 0.30 |  |  |
| *Nearest Road (inverse distance in meters)* |  |  |  |
| Mean (SD) | 0.07 (0.03) |  |  |
| Min, Max | 0.01, 0.20 |  |  |
| *Population Density (number of inhabitants per km<sup>2</sup>)* |  |  |  |
| Mean (SD) | 4857.80 (2756.99) |  |  |
| Min, Max | 1,000.00, 10,000.00 |  |  |
| *Green Space (Normalized Difference Vegetation Index, ranging from -1 to +1)* |  |  |  |
| Mean (SD) | 0.42 (0.09) |  |  |
| Min, Max | 0.20, 0.70 |  |  |
| *Walkability (average walkability index score (0-1))* |  |  |  |
| Mean (SD) | 0.30 (0.07) |  |  |
| Min, Max | 0.15, 0.50 |  |  |

| Building Density (no. of m<sup>2</sup> of buildings per km<sup>2</sup>) |  |  |  |
| --- | --- | --- | --- |
| Mean (SD) | 399458.73 (126787.83) |  |  |
| Min, Max | 20,000.00, 670,000.00 |  |  |
| Bus Lines (meters of bus lines in length per km<sup>2</sup>) |  |  |  |
| Mean (SD) | 3140.92 (1579.28) |  |  |
| Min, Max | 1,000.00, 8,000.00 |  |  |
| Connectivity Density (no. of road intersections per km<sup>2</sup>) |  |  |  |
| Mean (SD) | 129.34 (60.45) |  |  |
| Min, Max | 10.00, 340.00 |  |  |
| Depressive Symptoms |  |  |  |
| Mean (SD) | 4.46 (3.09) | 4.24 (3.25) | 14.66 (4.36) |
| Min, Max | 0.00, 16.00 | 0.00, 16.00 | 3.00, 25.00 |
| Contextual SES Risk |  |  |  |
| Mean (SD) | 1.69 (0.48) |  |  |
| Min, Max | 1.00, 4.25 |  |  |
| Smoking |  |  |  |
| Smoking | 19.58% |  |  |
| Former Smoker | 31.30% |  |  |
| Never smoked | 49.12% |  |  |
| Length of time at address (no. of years since last moved) |  |  |  |
| Mean (SD) | 3.01 (3.67) |  |  |
| Min, Max | 0.00, 34.00 |  |  |
| Familial History of Depression |  |  |  |
| Both parents | 2.09% |  |  |
| One parent | 21.40% |  |  |
| Neither parents | 76.51% |  |  |

Built environment features were intercorrelated (-0.66 to 0.96), as were depressive symptoms over time (0.04 to 0.42) (**Figure S1**). The strongest association among built environment features was between facility richness and facility density (*r* = 0.96, *p* < .0001). The strongest depressive symptoms association was between study enrolment and the 4-year follow-up (*r* = 0.42, *p* < .0001).

#### Overview of Network Analyses

In network 1, of 28 possible edges between built environment features and depressive symptoms at study enrolment, 23 showed nonzero associations with a mean edge weight of 0.03 (SD=0.18; range -0.31 to 0.58, within a possible range of -1.0 to 1.0; **Figure 1**; **Table 2**). In network 2, of 28 possible edges between built environment features and depressive symptoms at 4-year follow-up, 21 showed nonzero associations with a mean edge weight of 0.03 (SD=0.16; range -0.31 to 0.58; **Figure 1**; **Table 2**). In network 3, of 28 possible edges between built environment features and depressive symptoms at 18-year follow-up, 24 showed nonzero associations with a mean edge weight of 0.02 (SD=0.15; range -0.31 to 0.58; **Figure 1**; **Table 2**).

**Figure 1.**
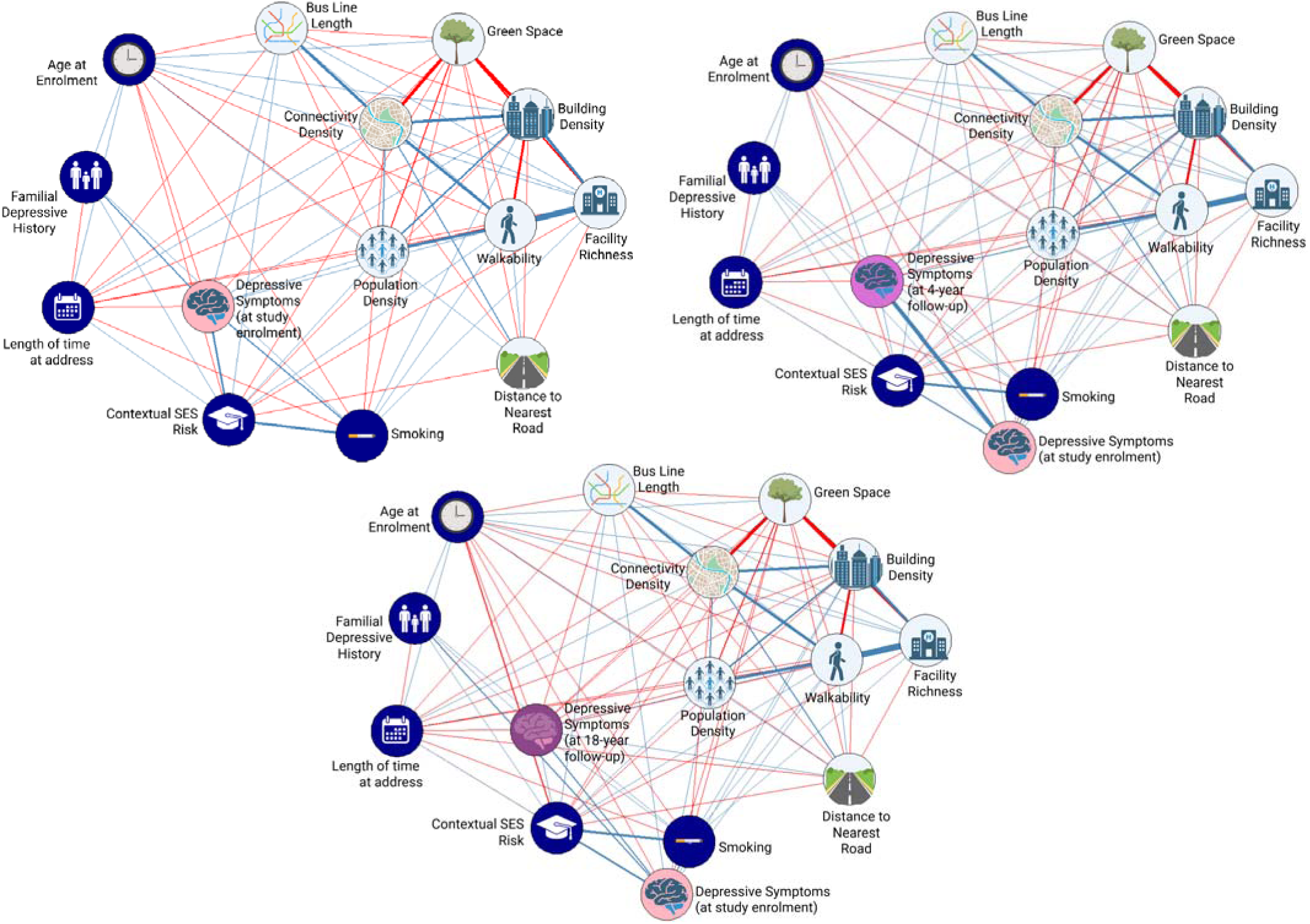
Network analysis of built environment features with depressive symptoms. Circles (nodes) represent variables (i.e., built environment measures or depressive symptoms) and lines (edges) represent the unique conditional (on all variables, including covariates) relationship between variables. Line thickness represents the strength of edges, and line colour (blue=positive, red=negative) represent the directionality of the edge. Top left panel) Depressive symptoms at study enrolment. Top right panel) Depressive symptoms at 4-year follow-up. Bottom panel) Depressive symptoms at 18-year follow-up.

**Table 2.**
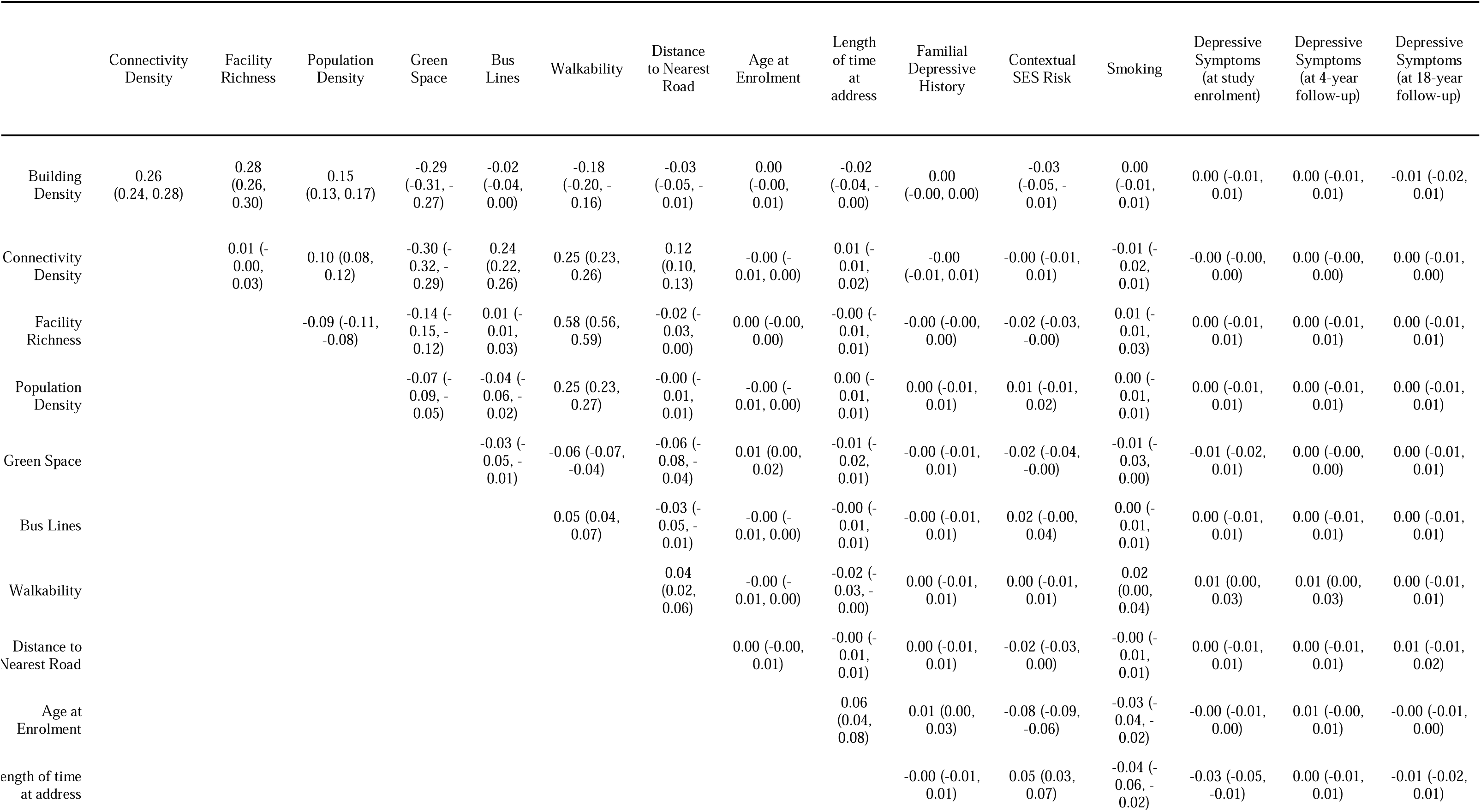
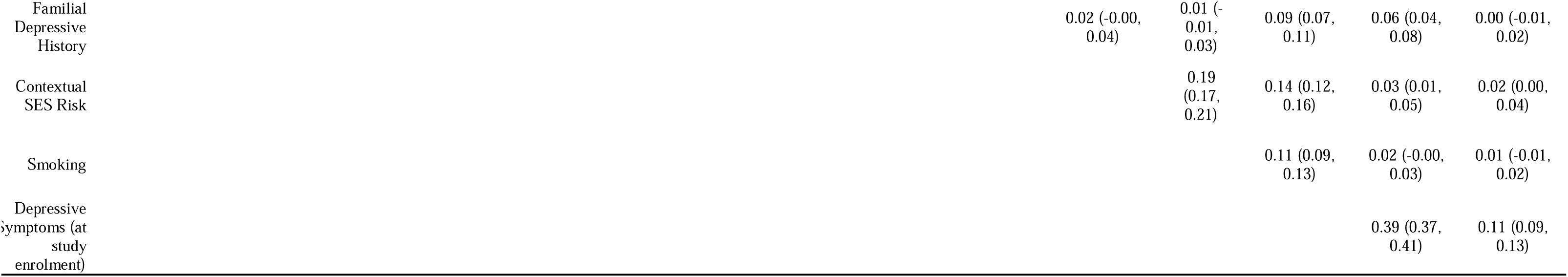
Pooled MICE edge weights across built environment features and depressive symptoms at study enrolment, 4-year follow-up and 18-year follow-up. Values in brackets represent bootstrapped 95% confidence intervals, reflecting the variability of edge weights across resampled datasets.

|  | Connectivity<br>Density | Facility<br>Richness | Population<br>Density | Green<br>Space | Bus<br>Lines | Walkability | Distance<br>to Nearest<br>Road | Age at<br>Enrolment | Length<br>of time<br>at<br>address | Familial<br>Depressive<br>History | Contextual<br>SES Risk | Smoking | Depressive<br>Symptoms<br>(at study<br>enrolment) | Depressive<br>Symptoms<br>(at 4-year<br>follow-up) | Depressive<br>Symptoms<br>(at 18-year<br>follow-up) |
| --- | --- | --- | --- | --- | --- | --- | --- | --- | --- | --- | --- | --- | --- | --- | --- |
| Building<br>Density | 0.26<br>(0.24, 0.28) | 0.28<br>(0.26, 0.30) | 0.15<br>(0.13, 0.17) | -0.29<br>(-0.31, -<br>0.27) | -0.02<br>(-0.04, 0.00) | -0.18<br>(-0.20, -<br>0.16) | -0.03<br>(-0.05, -<br>0.01) | 0.00<br>(-0.00, 0.01) | -0.02<br>(-0.04, -<br>0.00) | 0.00<br>(-0.00, 0.00) | -0.03<br>(-0.05, -<br>0.01) | 0.00<br>(-0.01, 0.01) | 0.00 (-0.01, 0.01) | 0.00 (-0.01, 0.01) | -0.01 (-0.02, 0.01) |
| Connectivity<br>Density |  | 0.01 (-<br>0.00, 0.03) | 0.10 (0.08, 0.12) | -0.30 (-<br>0.32, -<br>0.29) | 0.24<br>(0.22, 0.26) | 0.25 (0.23, 0.26) | 0.12<br>(0.10, 0.13) | -0.00 (-<br>0.01, 0.00) | 0.01 (-<br>0.01, 0.02) | -0.00<br>(-0.01, 0.01) | -0.00 (-0.01, 0.01) | -0.01 (-<br>0.02, 0.01) | -0.00 (-0.00, 0.00) | 0.00 (-0.00, 0.00) | 0.00 (-0.01, 0.00) |
| Facility<br>Richness |  |  | -0.09 (-0.11, -0.08) | -0.14 (-<br>0.15, -<br>0.12) | 0.01 (-<br>0.01, 0.03) | 0.58 (0.56, 0.59) | -0.02 (-<br>0.03, 0.00) | 0.00 (-0.00, 0.00) | -0.00 (-<br>0.01, 0.01) | -0.00 (-0.00, 0.00) | -0.02 (-0.03, -0.00) | 0.01 (-<br>0.01, 0.03) | 0.00 (-0.01, 0.01) | 0.00 (-0.01, 0.01) | 0.00 (-0.01, 0.01) |
| Population<br>Density |  |  |  | -0.07 (-<br>0.09, -<br>0.05) | -0.04 (-<br>0.06, -<br>0.02) | 0.25 (0.23, 0.27) | -0.00 (-<br>0.01, 0.01) | -0.00 (-<br>0.01, 0.00) | 0.00 (-<br>0.01, 0.01) | 0.00 (-0.01, 0.01) | 0.01 (-0.01, 0.02) | 0.00 (-<br>0.01, 0.01) | 0.00 (-0.01, 0.01) | 0.00 (-0.01, 0.01) | 0.00 (-0.01, 0.01) |
| Green Space |  |  |  |  | -0.03 (-<br>0.05, -<br>0.01) | -0.06 (-0.07, -0.04) | -0.06 (-<br>0.08, -<br>0.04) | 0.01 (0.00, 0.02) | -0.01 (-<br>0.02, 0.01) | -0.00 (-0.01, 0.01) | -0.02 (-0.04, -0.00) | -0.01 (-<br>0.03, 0.00) | -0.01 (-0.02, 0.01) | 0.00 (-0.00, 0.00) | 0.00 (-0.01, 0.01) |
| Bus Lines |  |  |  |  |  | 0.05 (0.04, 0.07) | -0.03 (-<br>0.05, -<br>0.01) | -0.00 (-<br>0.01, 0.00) | -0.00 (-<br>0.01, 0.01) | -0.00 (-0.01, 0.01) | 0.02 (-0.00, 0.04) | 0.00 (-<br>0.01, 0.01) | 0.00 (-0.01, 0.01) | 0.00 (-0.01, 0.01) | 0.00 (-0.01, 0.01) |
| Walkability |  |  |  |  |  |  | 0.04<br>(0.02, 0.06) | -0.00 (-<br>0.01, 0.00) | -0.02 (-<br>0.03, -<br>0.00) | 0.00 (-0.01, 0.01) | 0.00 (-0.01, 0.01) | 0.02<br>(0.00, 0.04) | 0.01 (0.00, 0.03) | 0.01 (0.00, 0.03) | 0.00 (-0.01, 0.01) |
| Distance to<br>Nearest Road |  |  |  |  |  |  |  | 0.00 (-0.00, 0.01) | -0.00 (-<br>0.01, 0.01) | 0.00 (-0.01, 0.01) | -0.02 (-0.03, 0.00) | -0.00 (-<br>0.01, 0.01) | 0.00 (-0.01, 0.01) | 0.00 (-0.01, 0.01) | 0.01 (-0.01, 0.02) |
| Age at<br>Enrolment |  |  |  |  |  |  |  |  | 0.06<br>(0.04, 0.08) | 0.01 (0.00, 0.03) | -0.08 (-0.09, -0.06) | -0.03 (-<br>0.04, -<br>0.02) | -0.00 (-0.01, 0.00) | 0.01 (-0.00, 0.01) | -0.00 (-0.01, 0.00) |
| Length of time<br>at address |  |  |  |  |  |  |  |  |  | -0.00 (-0.01, 0.01) | 0.05 (0.03, 0.07) | -0.04 (-<br>0.06, -<br>0.02) | -0.03 (-0.05, -0.01) | 0.00 (-0.01, 0.01) | -0.01 (-0.02, 0.01) |

|  |  |  |  |  |  |
| --- | --- | --- | --- | --- | --- |
| Familial<br>Depressive<br>History | 0.02 (-0.00,<br>0.04) | 0.01 (-<br>0.01,<br>0.03) | 0.09 (0.07,<br>0.11) | 0.06 (0.04,<br>0.08) | 0.00 (-0.01,<br>0.02) |
| Contextual<br>SES Risk |  | 0.19<br>(0.17,<br>0.21) | 0.14 (0.12,<br>0.16) | 0.03 (0.01,<br>0.05) | 0.02 (0.00,<br>0.04) |
| Smoking |  |  | 0.11 (0.09,<br>0.13) | 0.02 (-0.00,<br>0.03) | 0.01 (-0.01,<br>0.02) |
| Depressive<br>symptoms (at<br>study<br>enrolment) |  |  |  | 0.39 (0.37,<br>0.41) | 0.11 (0.09,<br>0.13) |

Nonparametric bootstrapping showed edge weights were determined with acceptable accuracy, evidenced by small variation across 1,000 bootstrap samples, with 95% confidence intervals (**Figures S2, S3, S4**). We identified the strongest edge (association) between facility richness and walkability (0.58) (**Table 2; Figure S2**).

Case-drop bootstrapping suggested measures of closeness and expected influence were stable across all three networks, with Coefficient of Stability (CS) values of 0.75 for Networks 1 and 2, and 0.73 for Network 3—exceeding recommended thresholds (CS > 0.50; ^20^) (**Figure S5**). Betweenness measures were lower (Network 1: CS = 0.19, Network 2: 0.22, Network 3: 0.21), indicating variables rarely served as intermediaries in any network.

#### Aim 1: How features of the built environment relate to each other

Generally, built environment features associated well with each other. The strongest edge observed in networks 1, 2 and 3 was between facility richness and walkability (0.58; **Table 2**).

Whilst most built environment features were positively associated, green space negatively associated with other built environment features. For example, as population density increased green space decreased (-0.29).

We expected population density to have the greatest expected influence. However, green space and walkability consistently showed the highest expected influence (Network 1: green space z = −2.63, walkability z = 1.26, **Figure S6**; Network 2: z = −2.76, 1.240, **Figure S7**; Network 3: z = −2.74, 1.35, **Figure S8, S9**). Building density had the highest betweenness centrality in all three networks (z = 2.15, 2.16, 2.16), indicating it played a key role in linking other nodes. When considering covariates, age at environment in networks 1 and 2, and age at depressive symptoms in network 3, had the second greatest expected influence (z=1.31, z=1.62 and z =1.37 respectively; **Table S1**).

#### Aim 2: Which specific features of the built environment have the strongest associations with depression

Overall, associations between built environment features and depressive symptoms were generally weak (all edge weights < ±0.02) and directions of association were unstable. At study enrolment, depressive symptoms had edges with walkability (0.01, CI: 0.00, 0.03) and green space (-0.01, CI: -0.02, 0.01). At the 4-year follow-up, as hypothesized an edge was observed between depressive symptoms and walkability (-0.01, CI: 0.00, 0.03). By the 18-year follow-up, weak edges between depressive symptoms and built environment variables included building density (-0.01, CI: -0.02, 0.01) and distance to the nearest road (0.01, CI: -0.01, 0.02).

#### Aim 3: How associations between built environment and depression change over time

Overall, unlike hypothesised, associations between built environment features and depressive symptoms were consistently weak with inconsistent effect sizes, likely reflecting estimation variability as opposed to meaningful change.

#### Exploratory Analysis: Do built environment features collectively predict depressive symptoms?

As there were no clear associations between individual built environment features and depressive symptoms, we conducted exploratory and confirmatory factor analyses. This was important to test as network analyses remove shared variance, meaning correlated built environment features may appear weak individually even if their combined relationship with depressive symptoms is more substantial ^20^.

Exploratory factor analysis (maximum likelihood extraction with oblimin rotation) indicated a dominant first factor (eigenvalue = 3.94), accounting for 49.2% of total variance (**SM Table S2**). All subsequent eigenvalues were below 1 (suggesting a steep drop-off after the first factor). Although parallel analysis suggested up to five factors, inspection of the scree plot and eigenvalue distribution indicated that a single general dimension captured the majority of shared variance.

In the one-factor solution, most indicators demonstrated moderate-to-strong loadings, although distance to the nearest road and bus line length loaded weakly (< .50) and were therefore removed from the reduced model. The retained factor accounted for 43.6% of variance in the full model and 54.6% following removal of weakly loading indicators.

A one-factor structure representing the built environment, comprising seven observed indicators (**Table 3**), was selected for parsimony, as similar model fits were observed across multi-factor models (**SM Table S3**; **Figure S10**). Global fit indices for this model were modest (CFI = 0.89, TLI = 0.68, RMSEA = 0.34, SRMR = 0.06), suggesting the current work may be more suitably interpreted as dimension reduction than reflecting a single latent construct.

**Table 3.** Factor loadings for one-factor model of built environment.

|  | Built Environment Factor |
| --- | --- |
| Building Density | 0.645 |
| Connectivity Density | 0.802 |
| Facility Richness | 0.809 |
| Green Space | -0.740 |
| Population Density | 0.512 |
| Bus Lines Length | 0.423 |
| Walkability | 0.870 |
| Distance to Nearest Road | 0.166 |

In regression analyses, the built environment factor significantly predicted depressive symptoms at age 28 (standardised β = 0.148, p < .001) and at age 32 (standardised β = 0.114, p = .011) controlling for age at enrolment, time at current address, familial history of depression, SES risk factors, smoking status and baseline depressive symptoms (only at age 32). However, the factor did not significantly predict depressive symptoms at the 18-year follow-up (**Table 4**).

**Table 4.** Regression of built environment factor with depressive symptoms at study enrolment, 4-year follow-up and an 18-year follow-up with covariates.

|  | Depressive Symptoms<br>(at study enrolment) |  | Depressive Symptoms<br>(at 4-year follow-up) |  | Depressive Symptoms<br>(at 18-year follow-up) |  |
| --- | --- | --- | --- | --- | --- | --- |
|  | Estimate | *p* value | Estimate | *p* value | Estimate | *p* value |
| Built Environment Factor | 0.148 (0.04) | < .001*** | 0.114 (0.04) | 0.011* | -0.005<br>(0.08) | 0.950 |
| Age (at enrolment) | -0.006 (0.05) | 0.912 | -0.031 (0.06) | 0.614 | 0.025 (0.10) | 0.802 |
| Familial Depressive History (at study enrolment) | 0.623 (0.06) | < .0001*** | 0.429 (0.08) | < .0001*** | 0.057 (0.13) | 0.669 |
| SES Risk Factors (at study enrolment) | 1.097 (0.07) | < .0001*** | 0.320 (0.08) | .0001** | 0.239 (0.15) | 0.122 |
| Length of Time at Address (at study enrolment) | 0.0001 (0.01) | 0.963 | -0.004 (0.01) | 0.673 | -0.01 (0.02) | 0.561 |
| Smoking (at study enrolment) | 0.481 (0.04) | < .0001*** | 0.116 (0.04) | 0.007 | 0.034 (0.08) | 0.122 |
| Depressive Symptoms (at study enrolment) | - | - | 0.412 (0.01) | < .0001*** | 0.156 (0.02) | < .0001*** |
| Adjusted R<sup>2</sup> | 0.075 |  | 0.179 |  | 0.016 |  |

*Note.* Values represent standardised estimates and values in brackets denote standard errors. \*\*\* $p < .0001$ , \*\* $p < .001$ , \* $p < .05$

## Discussion

This study aimed to investigate 1) how built environment features relate to each other; 2) which built environment features have the strongest associations with depression and 3) how these associations change over time. Our results showed 1) built environment variables generally associated with each other; 2) network analyses associations between built environment variables and depressive symptoms centered around zero; 3) these associations were consistently weak across time. Follow-up analyses showed a built environment factor modestly associated with depressive symptoms at study enrolment, and 4-year follow-up, but not at an 18-year follow-up. We highlight and discuss five key points.

Firstly, built environment variables generally associated positively with each other, except green space. The strongest edge within built environment variables was between walkability and facility richness, which is consistent with facility richness being a component constituting walkability ^15,25^. Green space negatively associated with density-related built environment variables including population density, building density and connectivity density which is consistent with research finding less dense cities have greater proportions of green space ^26^. As hypothesised, we also found two largely separate clusters within built environment nodes, one for transport-related and another for facility/density related variables. This indicates relationships between built environment variables are in line with current understanding of how they interact. We hypothesised population density would exert the greatest influence in the networks. However, green space and walkability consistently showed the greatest expected influence across networks, suggesting built environment features related to mobility and accessibility may play a greater role in shaping relationships between the built environment and depressive symptoms than population density.

Secondly, most associations between built environment variables and depressive symptoms centered around zero. Whilst there were some non-zero associations, these were generally weak. For example, green space only had a weakly negative association with depressive symptoms at study enrolment and 18-year follow-up. This is inconsistent with literature identifying a protective effect of green space for depressive symptoms ^27^. Similarly, walkability had very weak and inconsistent associations with depressive symptoms at study enrolment and 4-year follow-up. This is at odds with previous literature identifying walkability as a protective factor for depression ^8,9,28^. However, by only including one or two built environment measures ^29^, this may have inflated individual estimates in previous research. Therefore, rather than indicating weak relationships between the built environment and depressive symptoms, our small edge weights instead may reflect the complex, multifactorial nature of relationships between urban environments and mental health.

Our exploratory analyses support this argument, as the built environment factor significantly associated with depressive symptoms at study enrolment and the 4-year follow-up. Whilst this study aimed to dissect relationships between built environment features and depression, this evidence supports it may be the collective contribution of these built environment features that better predict depressive symptoms. This suggests policies considering the built environment as an interrelated system, targeting multiple system features, may be more appropriate for protecting population mental health than policies focused on one component.

Thirdly, unlike hypothesised, associations between built environment variables and depressive symptoms were consistently weak across time. However, the built environment factor significantly associated with depressive symptoms at study enrolment, and the 4-year follow-up, but not the 18-year follow-up, aligning more closely with what we hypothesised. This may indicate relationships between individual components of the built environment and depressive symptoms are weak, whilst relationships between built environment factors and depressive symptoms may be stronger cross-sectionally then they are longitudinally.

Our findings must be considered alongside limitations. Firstly, standardized effect estimates for relationship between the built environment factor and depressive symptoms were modest. However, modest effects can have clinically meaningful impacts when scaled up to populations ^30^, which is particularly important considering the proportion of the global population exposed to urban environments ^31^. In addition, whilst depressive symptoms at study enrolment and the 4-year follow-up were measured using the CCEI score, the 18-year follow-up used the SF-36 mental health survey. Despite similar trends across both measures ^32^, this may have contributed to weakened longitudinal associations between the built environment factor and depressive symptoms at the 18-year follow-up. Lastly, we conducted EFA and CFA on the same dataset and so further replication in independent samples is recommended ^33^.

To conclude, this study aimed to investigate relationships between built environment features and depressive symptoms over time. Results indicated feature-specific relationships appeared to be generally weak. A built environment factor associated more consistently with depressive symptoms cross-sectionally and a 4-year follow-up period. Our evidence suggests a combined influence of built environment features, not individual effects, on depressive symptoms. Hence, multi-dimensional urban planning and system-level interventions may have more promise in improving community mental health than targeting single features of the built environment. Future research may wish to further explore these pathways and enhance understanding of factors that may strengthen or attenuate them.

## Methods

### Participants

Participants were mothers from the Avon Longitudinal Study of Parents and Children (ALSPAC) cohort ^34,35^. Pregnant women resident in Avon, UK with expected dates of delivery 1st April 1991 to 31st December 1992 were invited to take part in the study and the initial number of pregnancies enrolled was 14,541. We selected mothers who had at least 50% data available for built environment features (including building density (300m buffer), connectivity density (300m buffer), facility density (300m buffer), facility richness (300m buffer), bus lines length (300m buffer), bus stops (500m buffer), total traffic load (100m buffer), distance to nearest road, walkability (300m buffer), green space (500m buffer) and population density) and depressive symptoms at study enrolment, leaving up to 10,310 mothers available for analysis. We intended to also include bus stops and traffic load as built environment features, but this was not possible due to missing data (99.5% and 91.5% respectively). The study website (https://www.bristol.ac.uk/alspac/researchers/our-data/) contains details of all data available through a fully searchable data dictionary and variable search tool.

Ethical approval for the study was obtained from the ALSPAC Ethics and Law Committee and the Local Research Ethics Committees and Department of Psychology at the University of Bath. The study website (https://www.bristol.ac.uk/alspac/researchers/our-data/) contains details of all data available through a fully searchable data dictionary and variable search tool.

### Measures

#### Built Environment

All built environment variables used were derived through the Horizon 2020 LifeCycle project ^36,37^ and were obtained for participants’ addresses during the year of study enrolment using Geographic Information System (GIS; ^38^) software. Participant residential addresses were geo-coded to 1 m resolution ^39^. Built environment variables selected included building density, connectivity density, facility density, facility richness, bus lines length, distance to the nearest road, walkability, green space and population density (see **Figure 2**). For information on how each of these variables were defined, please see **supplementary section 1.1**.

**Figure 2.**
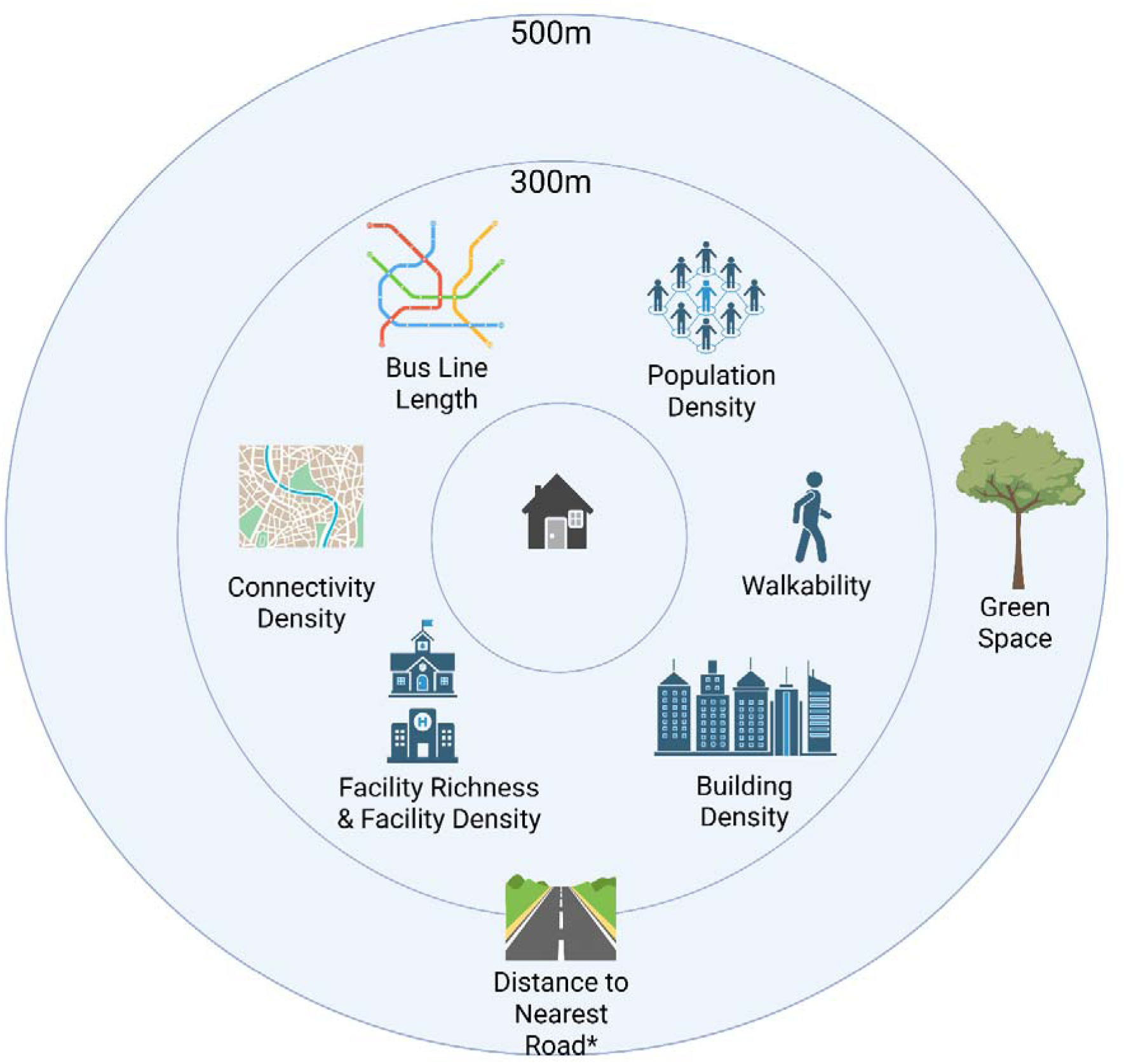
Built environment variables measured in this study and the distance buffers at which they were measured at. *Distance to nearest road did not have a distance buffer associated with it and instead was measured through absolute distance.

#### Depressive Symptoms

Depressive symptoms were measured at study enrolment and at a 4-year follow-up using the Crown Crisp Experiential Index (CCEI) ^40^. The CCEI consisted of 10 items such as, ‘I have felt sad or miserable’ and ‘I have been so unhappy that I have been crying’, with responses including ‘Yes, most of the time’, ‘Yes, quite often’, ‘Not very often/Only occasionally’ and ‘No, not at all/No, never’. Scores can range from 0 to 30. Depressive symptoms were also measured at an 18-year follow-up, using the 36-Item Short Form Survey (SF-36) Mental Health subscale, a valid and reliable measure of mental health ^41^ and a strong predictor of depression diagnosis ^42^.

#### Covariates

Covariates included contextual socioeconomic status (SES) risk factors, length of time at address, familial depressive history, age at built environment and depressive symptoms, and smoking (**supplementary section 1.2**).

### Statistical Analysis

This study was preregistered prior to analysis on the Open Science Framework (https://osf.io/mg7z6) with deviation from the preregistration clearly highlighted. Missing data was imputed. See **supplementary section 1.3** for further information and for a comparison between original and imputed data (**Supplementary Table S4**). Network analyses were conducted on pooled MICE data, rather than on a single averaged dataset. Correlations between variables were assessed and facility density and richness were highly collinear (*r* = 0.96, *p* < .0001). As a result, one had to be pruned to avoid node redundancy and unstable estimations in EBICglasso ^43^. The variable with the smallest association with depressive symptoms at study enrolment (facility density) was pruned.

Three weighted, nondirectional networks were created for each depressive symptom timepoint when investigating their relationship with built environment features at study enrolment. Networks were adjusted for covariates, including age at built environment measurement, age at depressive symptom measurement, length of time at address, familial depressive history, contextual SES risk, smoking and additionally baseline depressive symptoms in the two longitudinal networks (see **Figure 3**).

**Figure 3.**
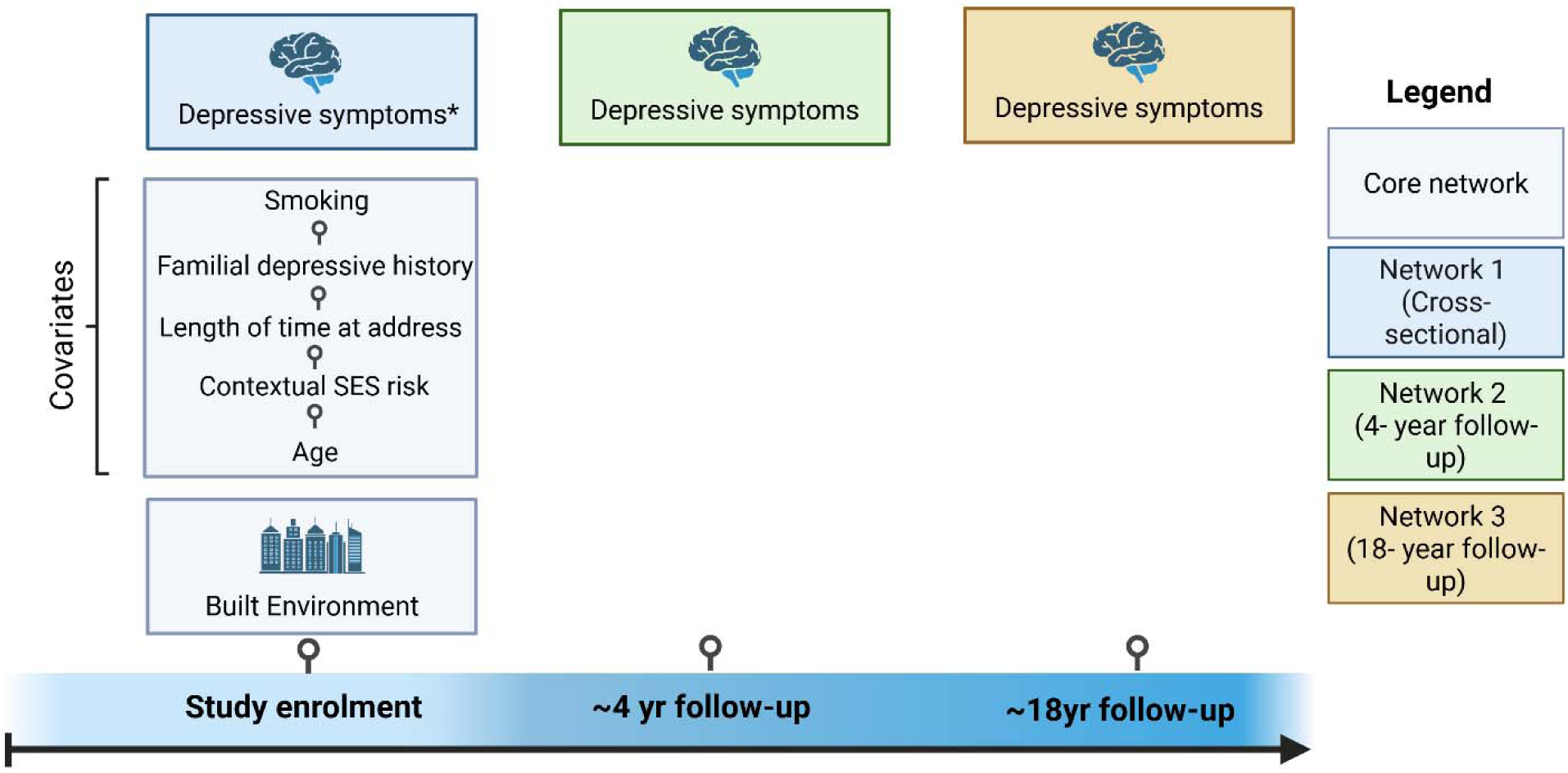
Analysis plan showing the timelines of variables. Core network refers to variables used across all networks. *Depressive symptoms at baseline are also used as a covariate in networks 2 and 3.

To perform network analysis, we used the qgraph package ^44^ in R Studio Statistical Software (v4.1.0; R Core Team, 2021), which uses a conservative EBICglasso and Gaussian Graphical Models using partial coefficients. For further details, please see **supplementary section 1.4**.

We performed nonparametric bootstrapping (1,000 resamples), with 95% confidence intervals (CIs), to assess accuracy of the network’s edge weights.

To understand how built environment features associate with each other (aim 1), we compared closeness and expected influence values for built environment variables. Closeness refers to the inverse of the total shortest-path distance from the node to all other nodes ^45^.

Expected influence refers to the sum of all edge weights connected to a node ^46^. To understand which built environment features have the strongest associations with depressive symptoms (aim 2), we compared edge weights between built environment variables and depressive symptoms. To test how associations change over time (aim 3), we compared the networks using depressive symptoms at study enrolment, 4 and 18-year follow-up.

As an exploratory (not preregistered) analysis, we also tested whether built environment features collectively were able to better predict depressive symptoms than built environment features on their own. To test this, we first performed an exploratory factor analysis (EFA) followed by a confirmatory factor analyses (CFA). The EFA and CFAs were conducted on the same dataset, due to the exploratory nature of this analysis and due to our large sample size ^33^.

The EFA was conducted in the psych package^47^ using maximum likelihood extraction with oblimin rotation, allowing factors to correlate. The number of factors were determined using parallel analysis and inspection of scree plots. Factor loadings ≥ .30 were considered meaningful, and variables with loadings < .50 in the one-factor solution were considered for removal. CFA models were then estimated in the lavaan package^48^ using robust maximum likelihood estimation (MLR), and model fit was evaluated using CFI, TLI, RMSEA, and SRMR.

Next, we conducted a series of multiple regressions testing whether the built environment factor predicted depressive symptoms at study enrolment, a 4-year follow up and an 18-year follow-up, whilst controlling for a number of covariates, including age at built environment measurement, age at depressive symptom measurement, length of time at address, familial depressive history, contextual SES risk, baseline depressive symptoms (for longitudinal models only) and smoking. For further information, see **supplementary section 1.4**.

## Data Availability

The study website (https://www.bristol.ac.uk/alspac/researchers/our-data/) contains details of all data available through a fully searchable data dictionary and variable search tool.

## Code availability

The code used to perform the analyses in this study is publicly available on GitHub at: https://github.com/fsanders1/built-environment-network.

## Acknowledgements

We are extremely grateful to all the families who took part in this study, the midwives for their help in recruiting them, and the whole ALSPAC team, which includes interviewers, computer and laboratory technicians, clerical workers, research scientists, volunteers, managers, receptionists and nurses.

## Ethics and consent statement

As described elsewhere^49^, ethical approval for the study was obtained from the ALSPAC Ethics and Law Committee and the Local Research Ethics Committees. Informed consent for the use of data collected via questionnaires and clinics was obtained from participants following the recommendations of the ALSPAC Ethics and Law Committee at the time. Informed consent was obtained from all subjects and/or their legal guardian(s). Participants can contact the study team at any time to retrospectively withdraw consent for their data to be used. Study participation is voluntary and during all data collection sweeps, information was provided on the intended use of data. Full details of the ALSPAC consent procedures are available on the study website (https://www.bristol.ac.uk/alspac/researchers/research-ethics/). The study website (https://www.bristol.ac.uk/alspac/researchers/our-data/) contains details of all data available through a fully searchable data dictionary and variable search tool.

