## supplemental materials for "A Network Analysis of Built Environment Features and Depressive Symptoms over an 18-year period"

**Figure S1.**

*Pooled correlation matrix (using imputed data) between all variables analysed in this study (facility density was later removed due to collinearity with facility richness).*

*^1^ Depressive symptoms measured at enrolment and a 4-year follow-up used the Crown Crisp Experiential Index. Depressive symptoms measured at ~18-year follow-up used the 36-Item Short Form Survey mental health scale.*

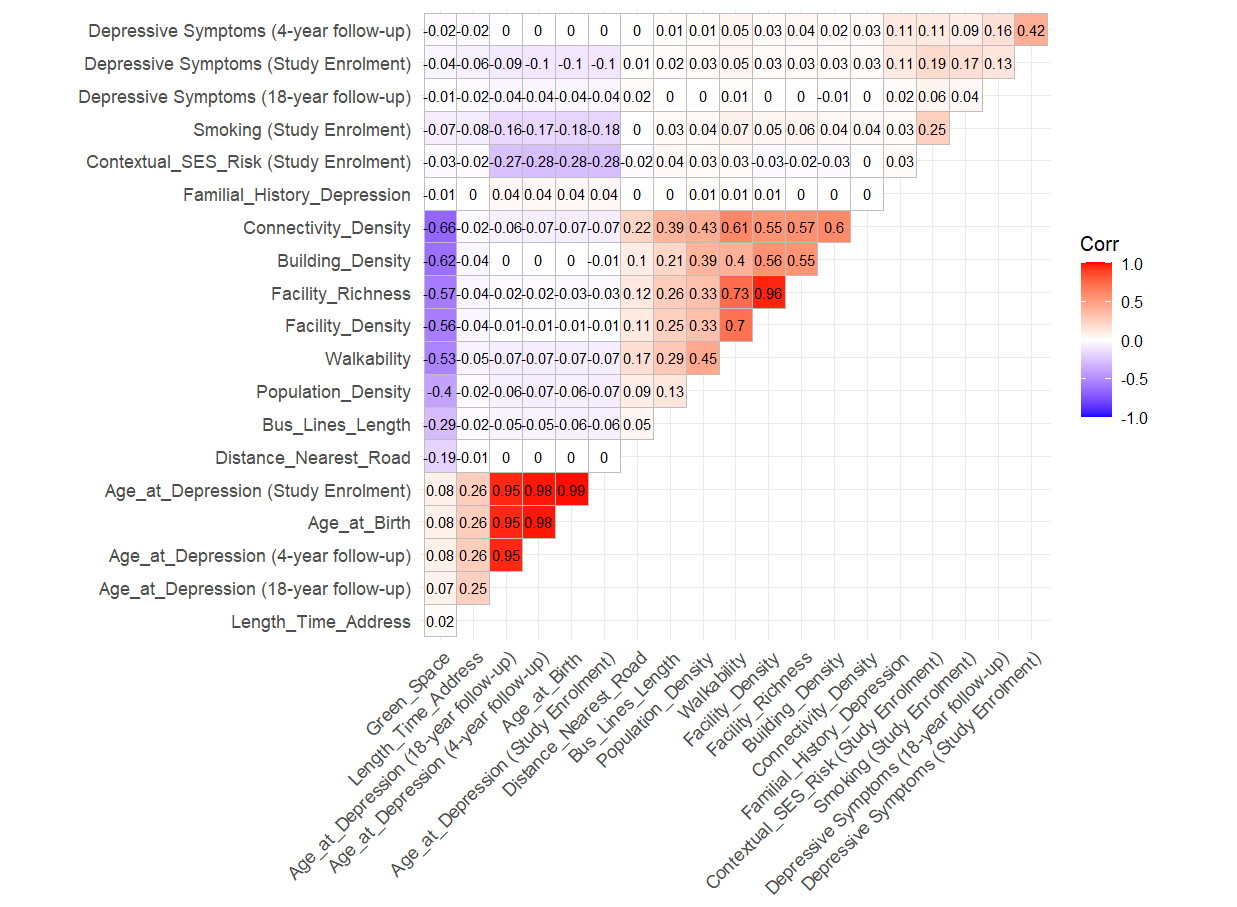

**Figure S2**

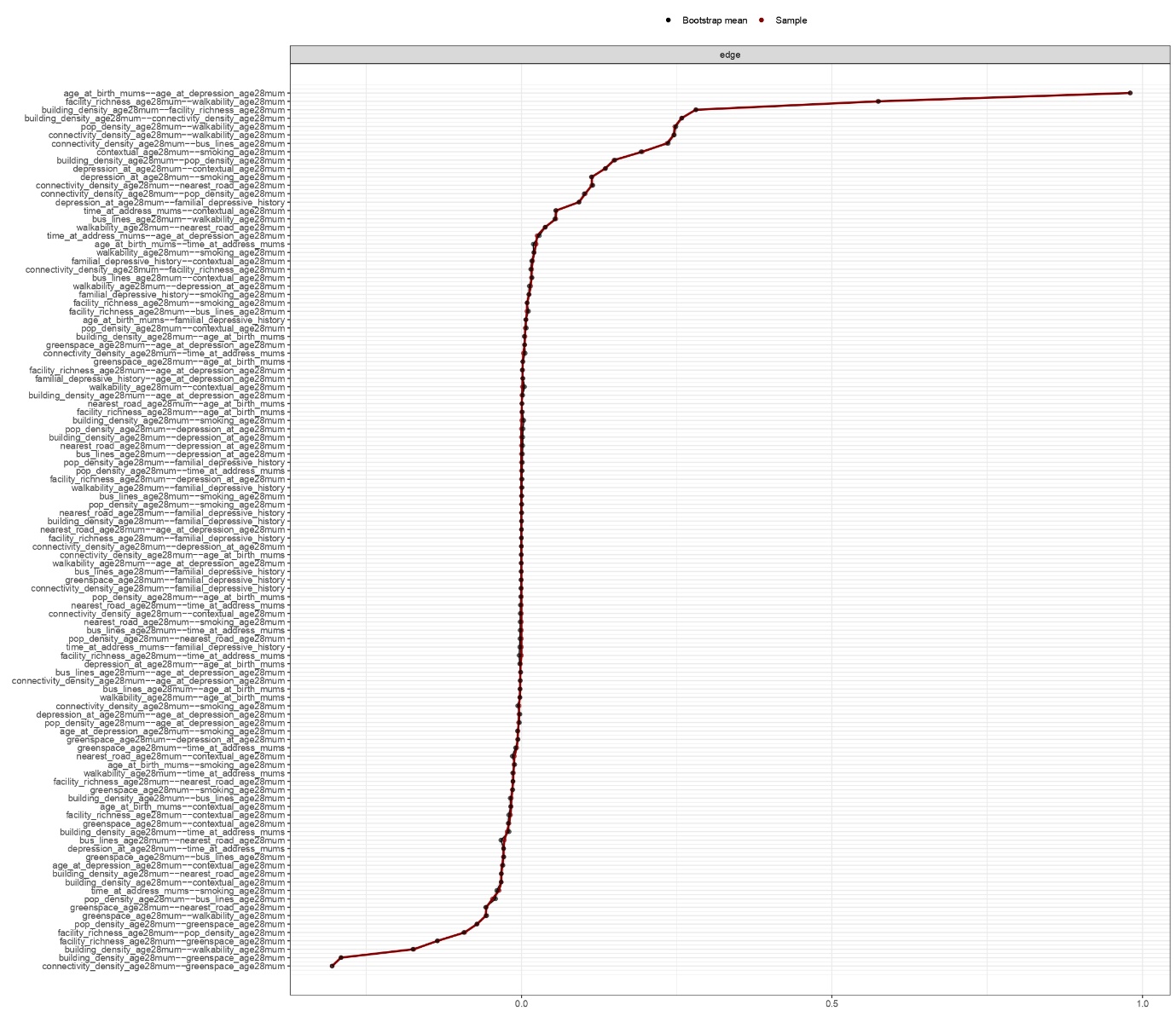
*Estimated edge weight accuracy in network 1 assessed through non-parametric bootstrapping (1,000 resamples). The sample edge weights are shown in red and the bootstrapped edge weights are shown in black.*

**Figure S3**

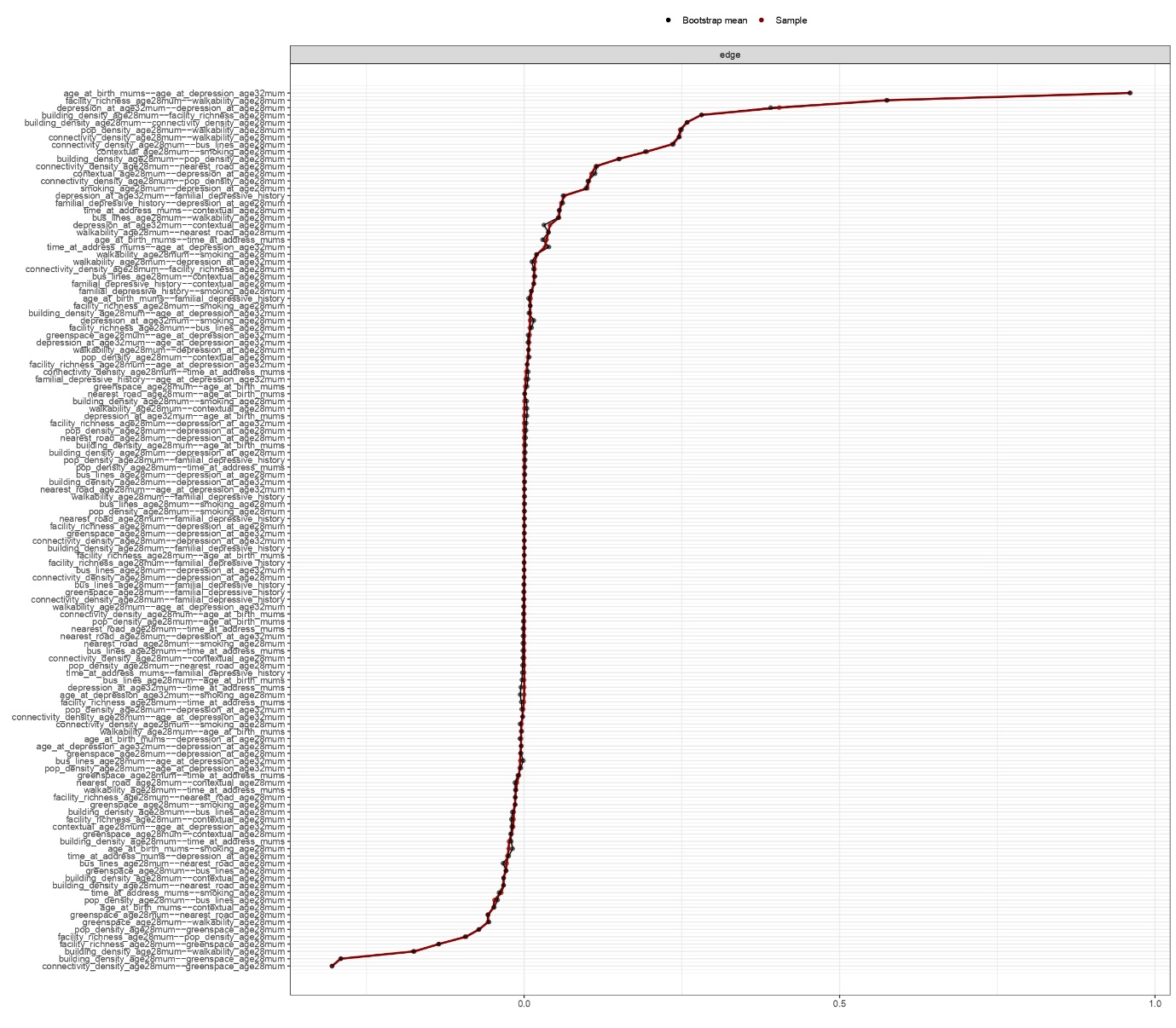
*Estimated edge weight accuracy in network 2 assessed through non-parametric bootstrapping (1,000 resamples). The sample edge weights are shown in red and the bootstrapped edge weights are shown in black.*

**Figure S4**

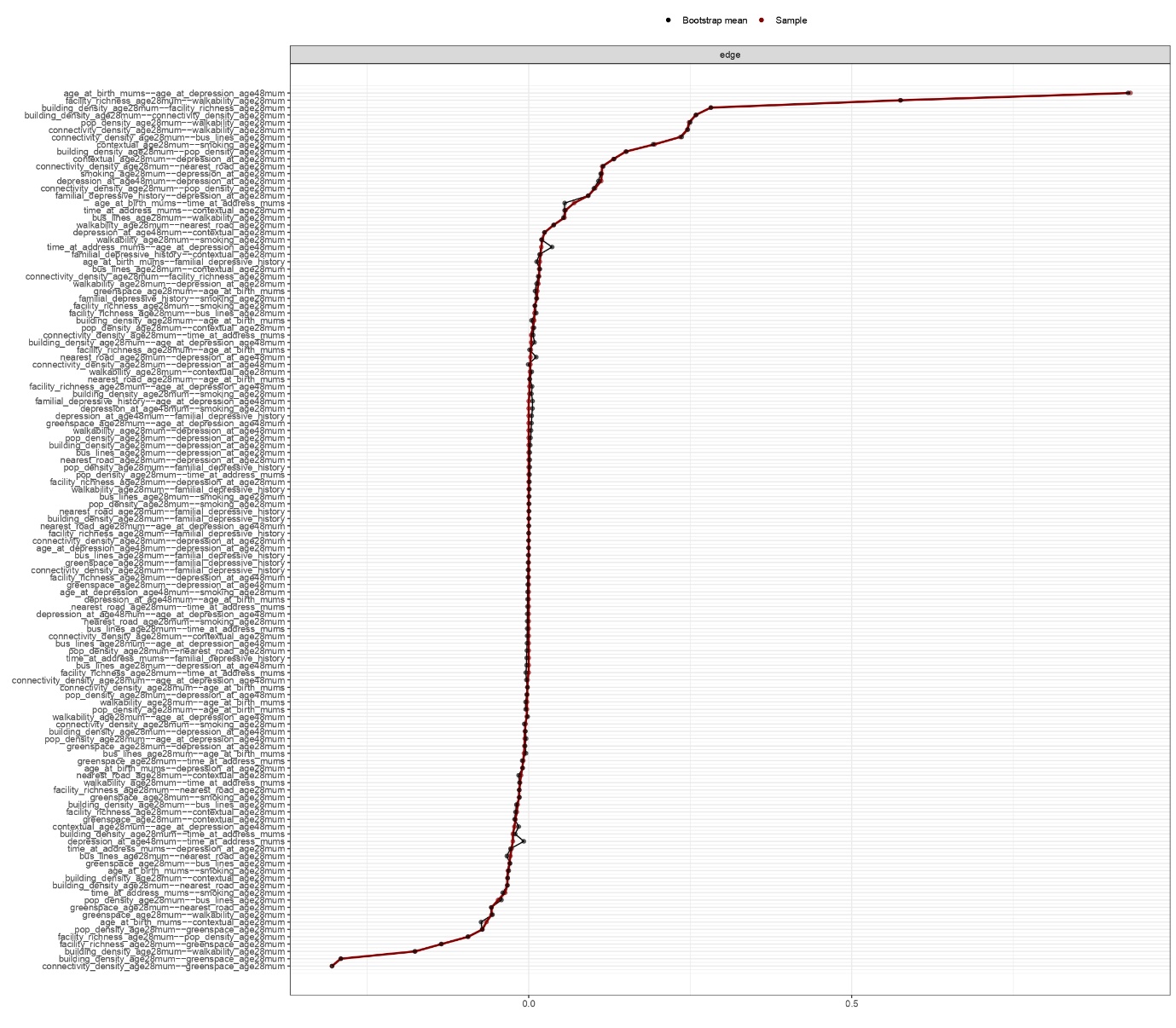
*Estimated edge weight accuracy in network 3 assessed through non-parametric bootstrapping (1,000 resamples). The sample edge weights are shown in red and the bootstrapped edge weights are shown in black.*

**Figure S5**

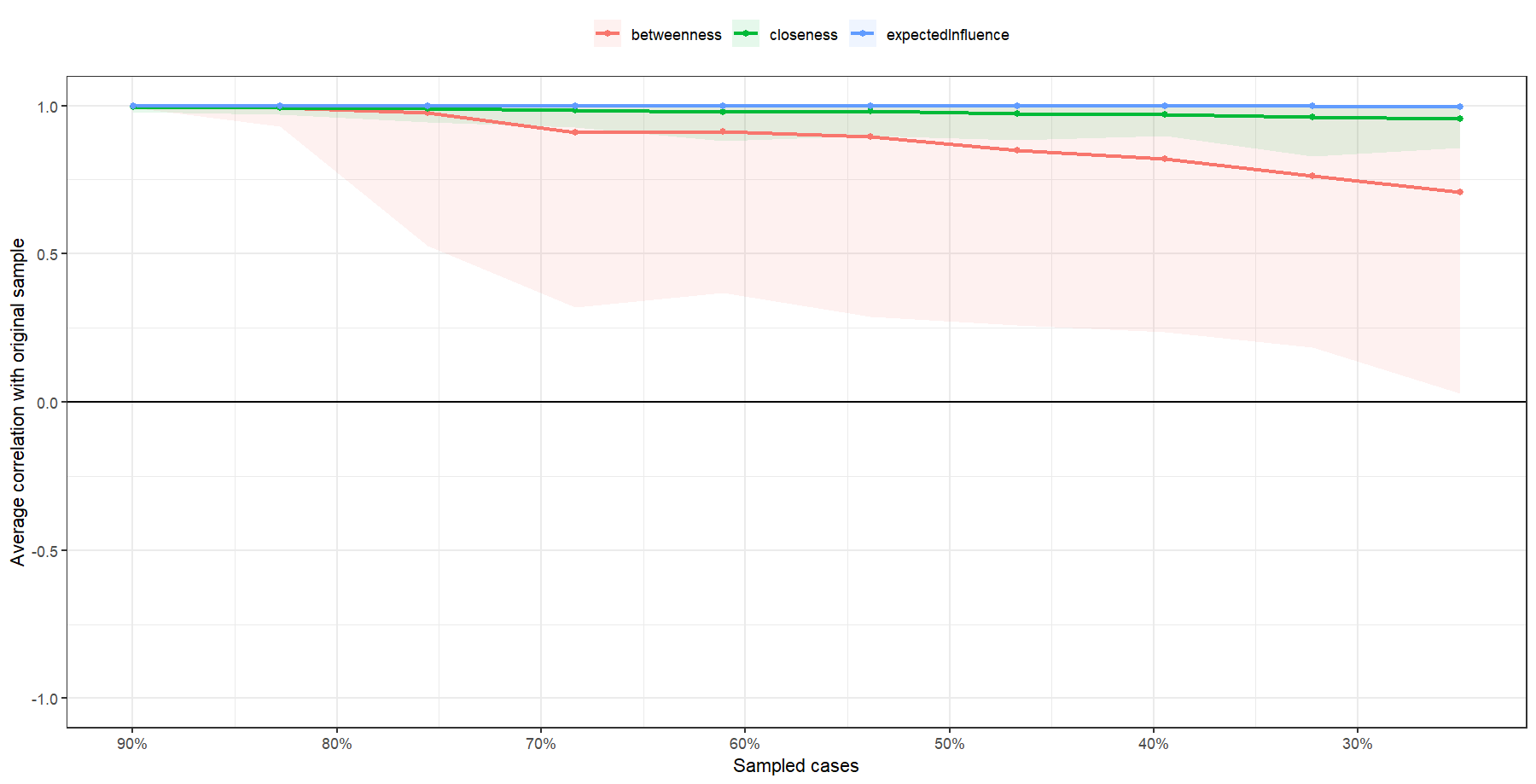

A

B
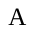

C
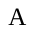

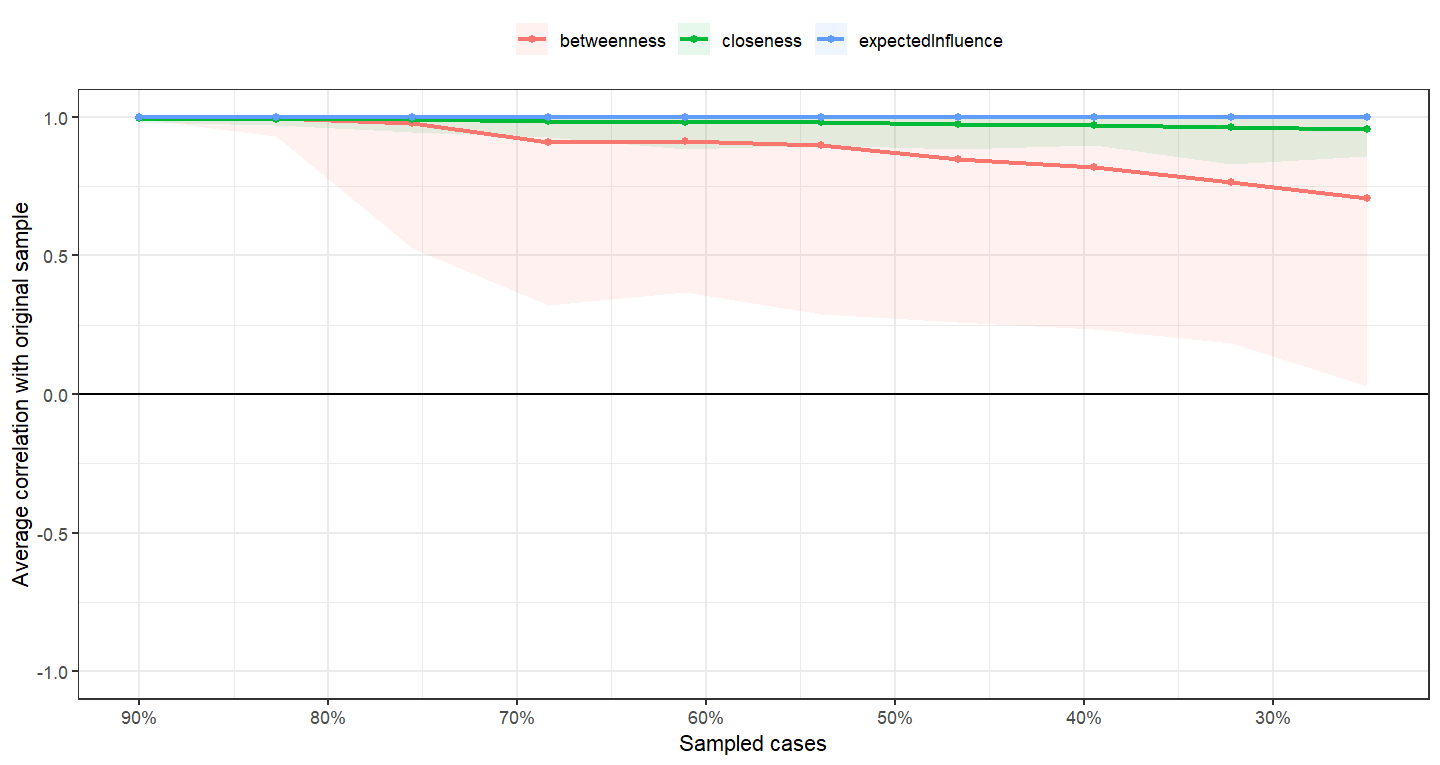

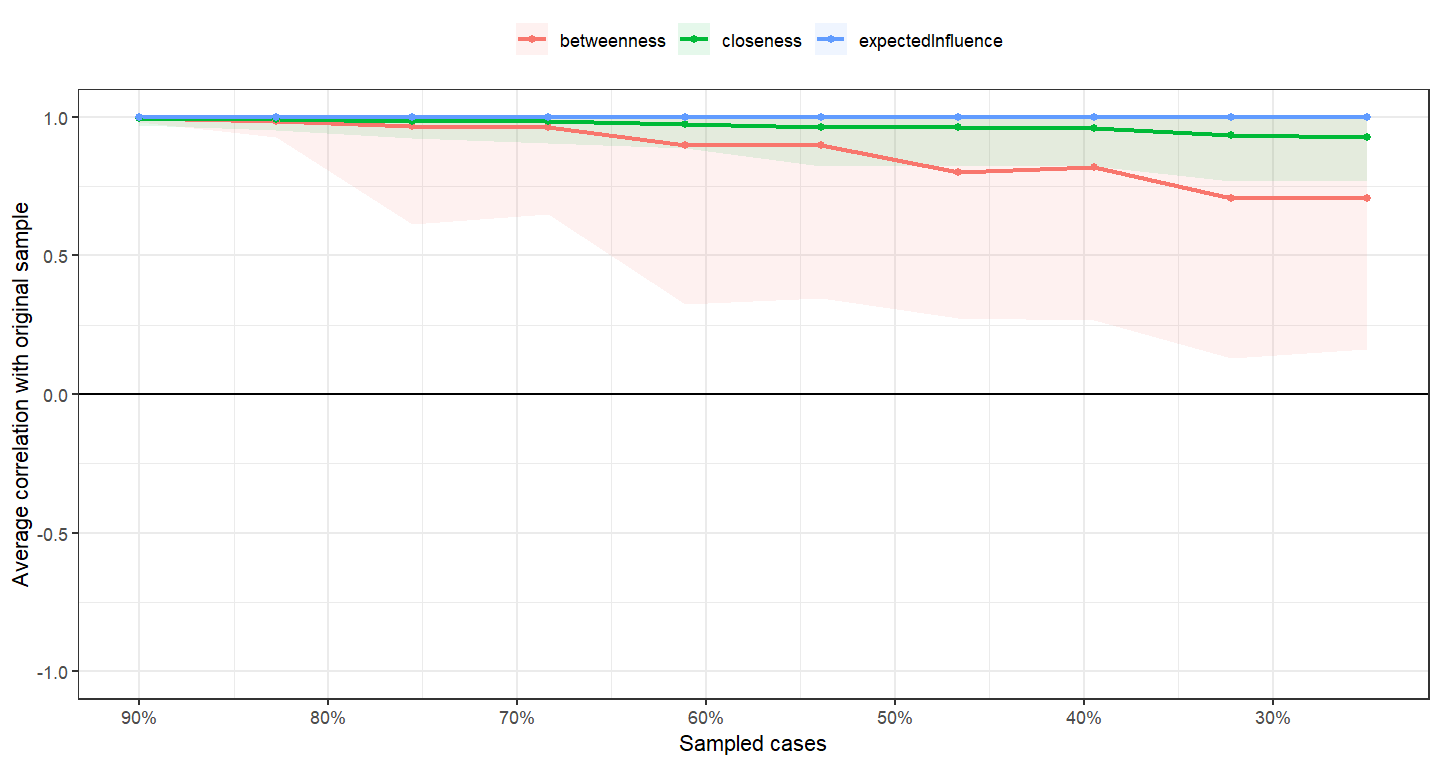

*Note.* These plots show the centrality stability assessed through case-drop bootstrapping (1,000 resamples) down to 20% of the original sample. The lines represent the correlations of centrality scores (e.g., betweenness, closeness, and expected influence) with the original sample and 95% confidence intervals are represented by the shaded areas.

**Figure S6**

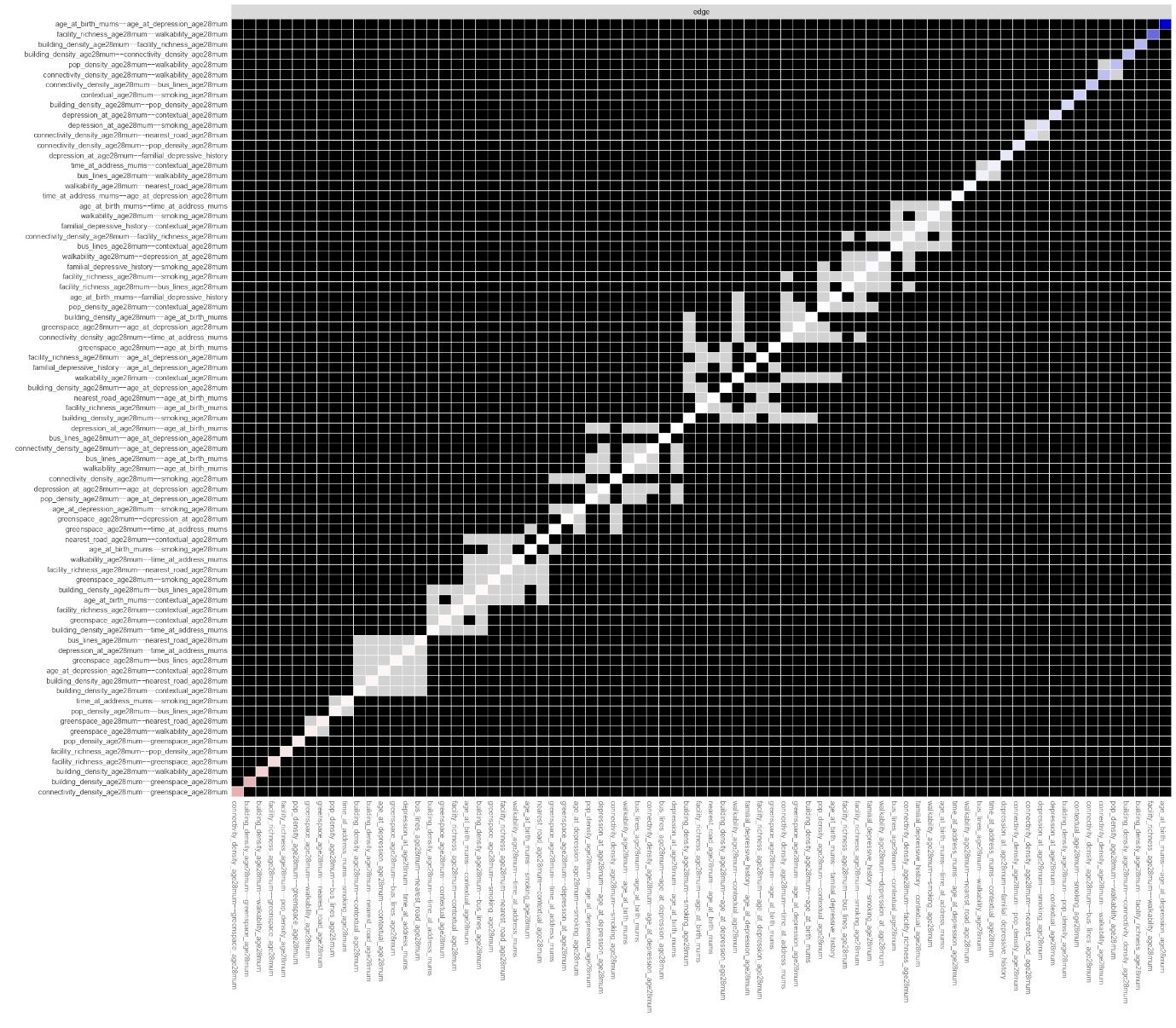
*Plot of edge weight difference test in network 1 using 95% confidence interval bootstrapping with 1,000 resamples. Black squares represent significant differences between the edge weights, and grey squares represent no significant differences. Significant differences are tested by comparing the observed edge weights to the distribution of bootstrapped values. Black squares represent the most robust edges in the network. The edges with the strongest associations are blue, with weakest associations in pink.*

**Figure S7**

*Plot of edge weight difference test in network 2 using 95% confidence interval bootstrapping with 1,000 resamples. Black squares represent significant differences between the edge weights, and grey squares represent no significant differences. Significant differences are tested by comparing the observed edge weights to the distribution of bootstrapped values. Black squares represent the most robust edges in the network. The edges with the strongest associations are blue, with weakest associations in pink.*

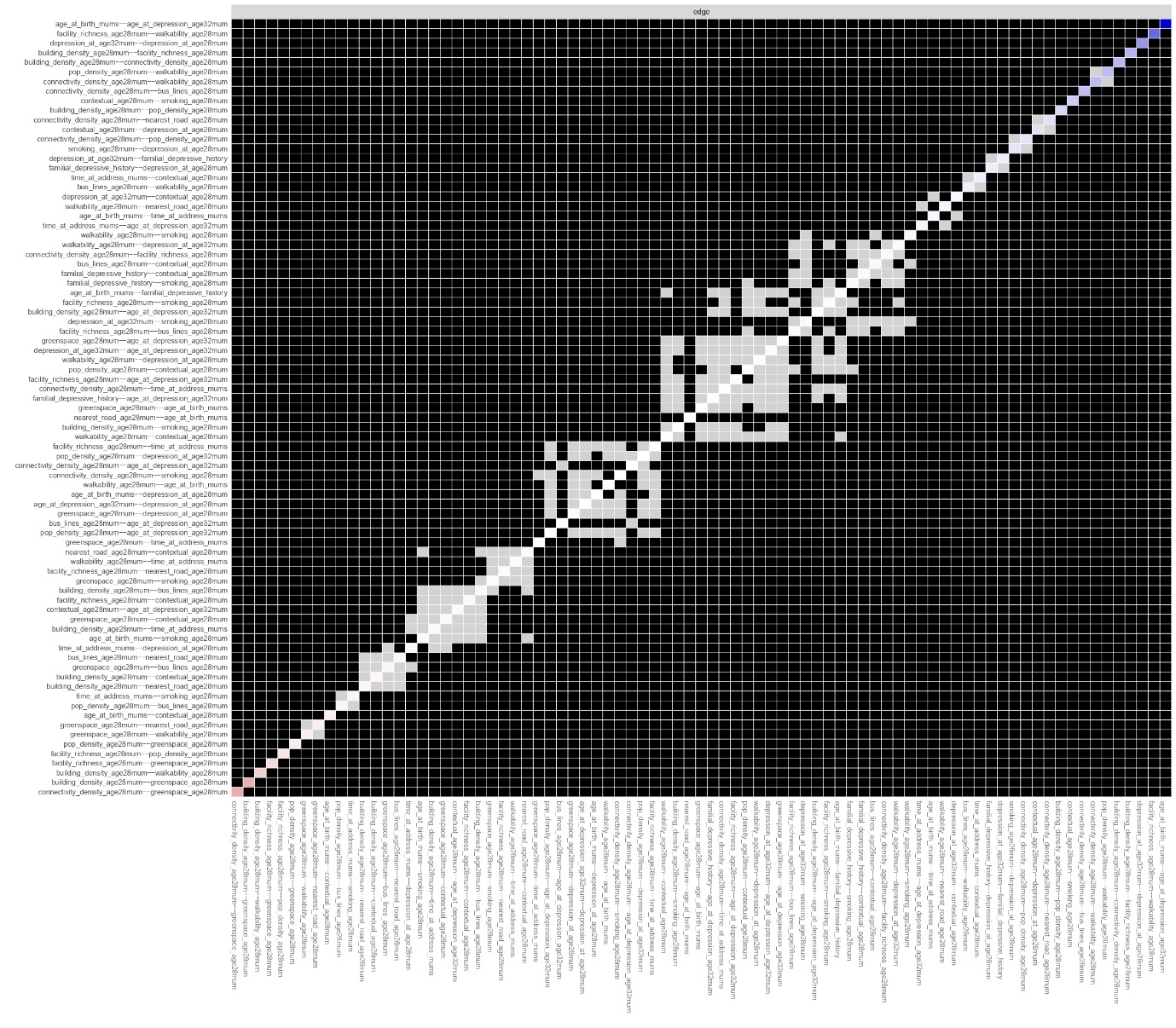

**Figure S8**

*Plot of edge weight difference test in network 3 using 95% confidence interval bootstrapping with 1,000 resamples. Black squares represent significant differences between the edge weights, and grey squares represent no significant differences. Significant differences are tested by comparing the observed edge weights to the distribution of bootstrapped values. Black squares represent the most robust edges in the network. The edges with the strongest associations are blue, with weakest associations in pink.*

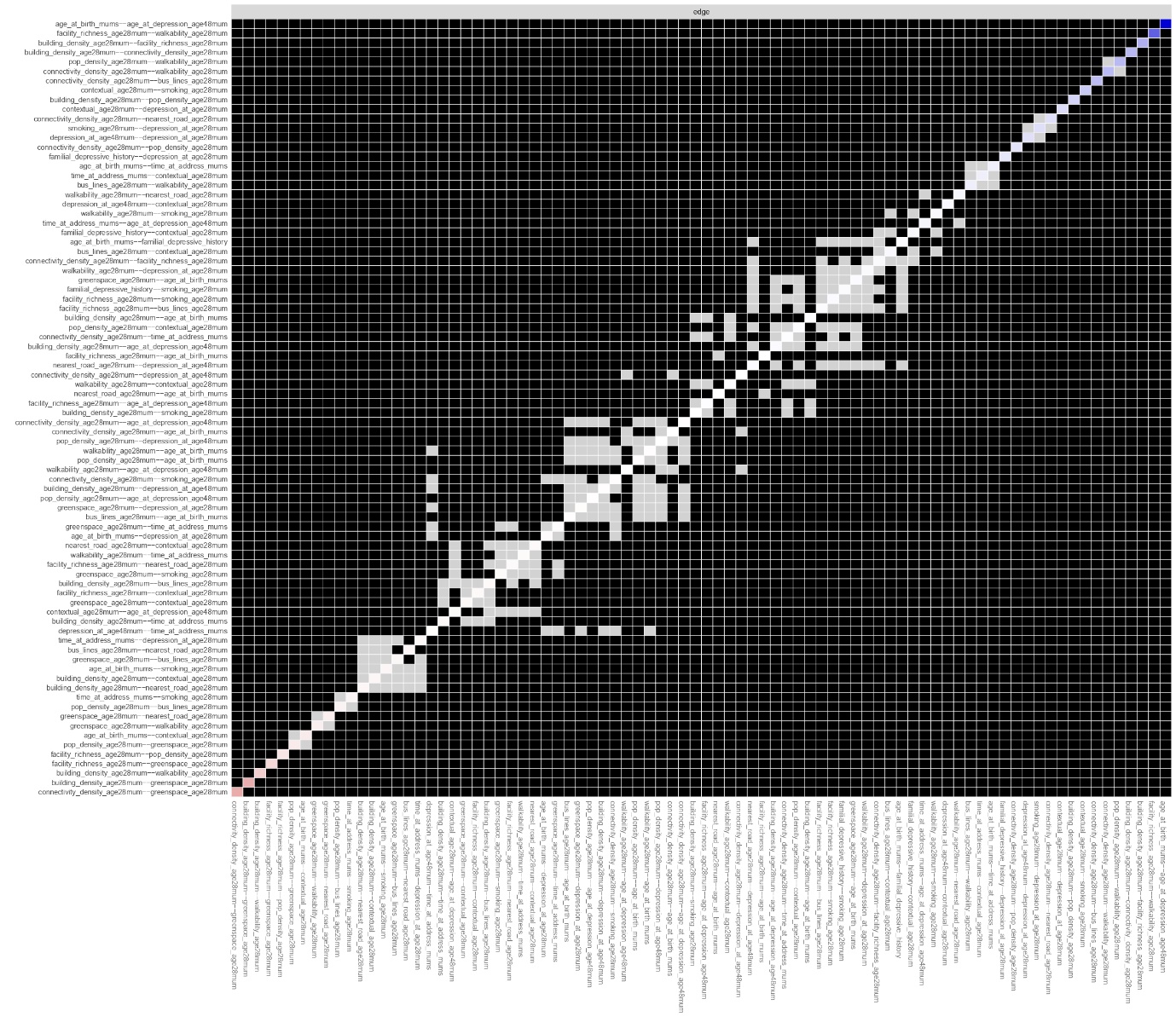

**Figure S9**

*Expected Influence plots. A: Network 1. B: Network 2. C: Network 3.*

A

B
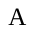

C
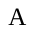

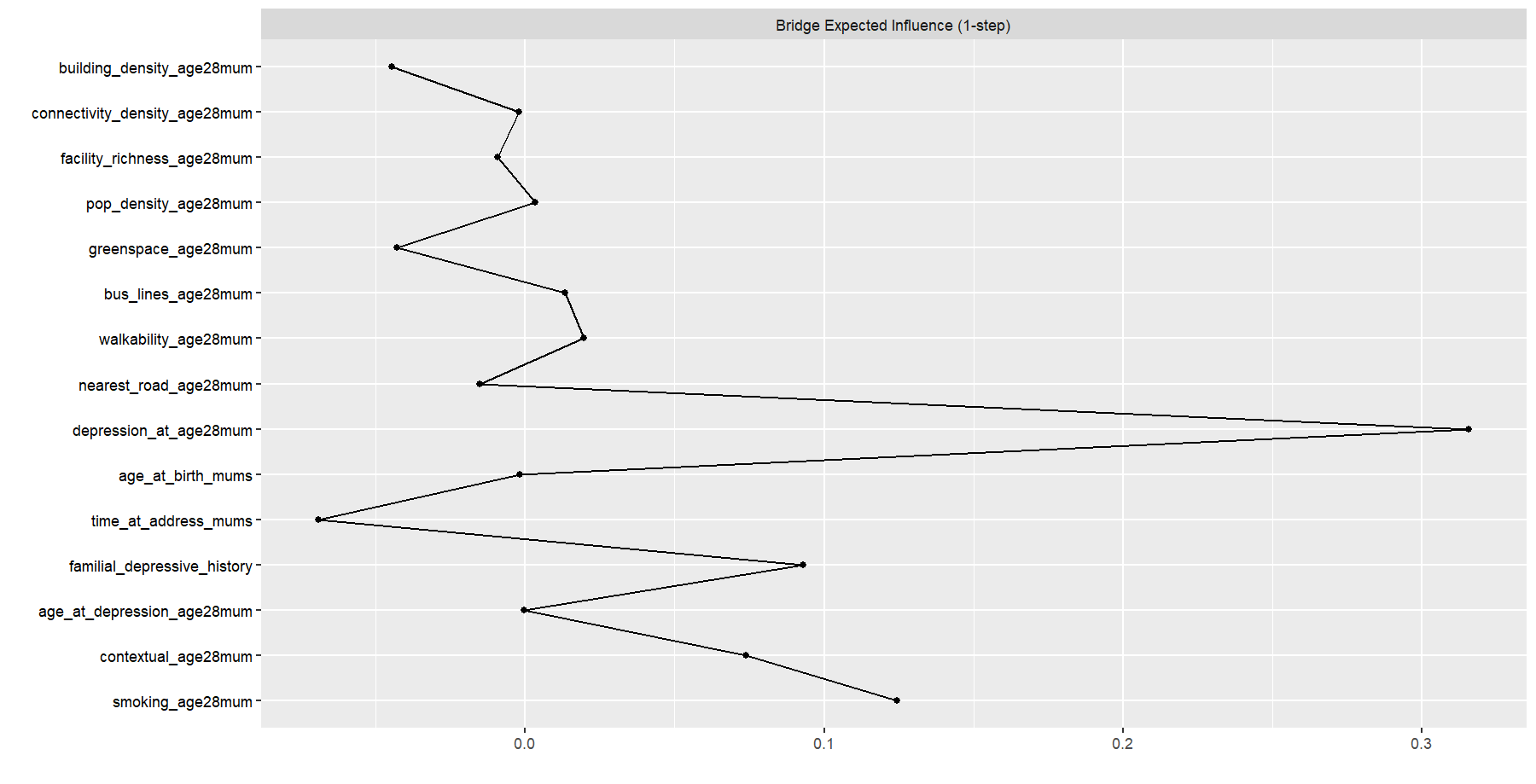

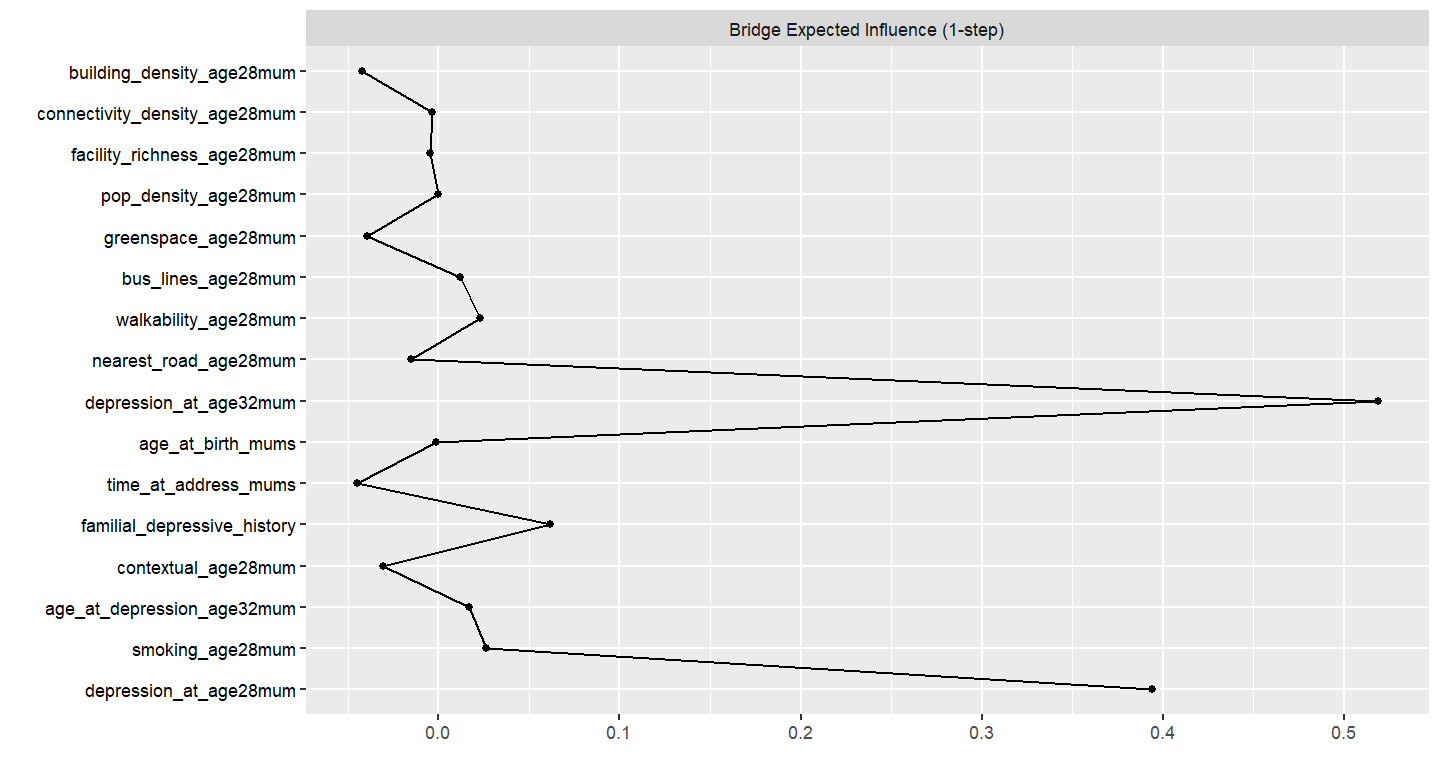

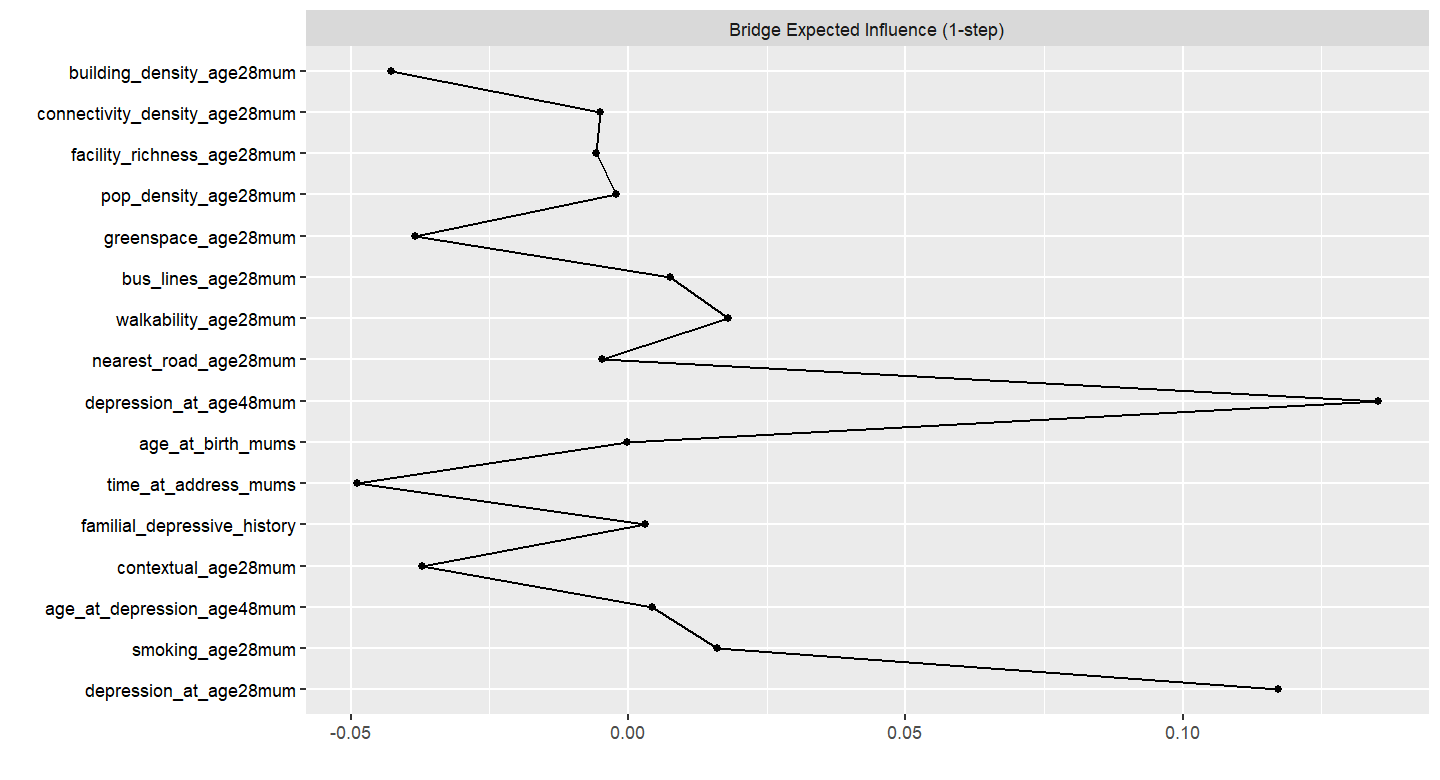

**Table S1.**

*Pooled centrality table for all networks.*

|  | Network 1 | | | Network 2 | | | Network 3 | | |
| --- | --- | --- | --- | --- | --- | --- | --- | --- | --- |
|  | Betweenness | Closeness | Expected Influence (1-step) | Betweenness | Closeness | Expected Influence (1-step) | Betweenness | Closeness | Expected Influence (1-step) |
| Building Density | 2.147 | 1.272 | -0.386 | 2.163 | 1.338 | -0.457 | 2.159 | 1.457 | -0.381 |
| Bus Lines Length | -0.627 | 0.259 | -0.258 | -0.622 | 0.101 | -0.335 | -0.618 | 0.011 | -0.264 |
| Connectivity Density | 0.794 | 0.952 | 0.686 | 0.741 | 0.920 | 0.639 | 0.722 | 0.964 | 0.746 |
| Facility Richness | -0.194 | 0.833 | 0.620 | -0.197 | 0.796 | 0.581 | -0.187 | 0.817 | 0.684 |
| Green Space | -0.627 | 0.811 | -2.625 | -0.622 | -1.780 | -2.760 | -0.618 | 0.789 | -2.740 |
| Distance to Nearest Road | -0.627 | -0.312 | -0.645 | -0.622 | 0.770 | -0.729 | -0.618 | -0.740 | -0.637 |
| Population Density | -0.627 | 0.372 | 0.050 | -0.622 | -0.571 | -0.124 | -0.618 | 0.181 | -0.041 |
| Walkability | -0.235 | 0.697 | 1.261 | -0.257 | -1.124 | 1.240 | -0.271 | 0.612 | 1.350 |
| Depressive Symptoms | 0.083 | -0.082 | -0.013 | -0.563 | -0.292 | 0.339 | -0.618 | -1.320 | -0.371 |
| Age at Environment | -0.627 | -1.815 | 1.314 | 0.022 | -1.349 | 1.168 | 0.050 | -0.821 | 1.205 |
| Age at Depressive Symptoms | 0.083 | -1.764 | 1.306 | -0.589 | -1.440 | 1.318 | -0.612 | -0.976 | 1.369 |
| Smoking | -0.627 | 0.093 | -0.102 | -0.622 | 0.249 | -0.124 | -0.618 | 0.316 | -0.108 |
| Length of time at address | -0.627 | -0.994 | -0.659 | -0.622 | 0.375 | -0.188 | -0.618 | -1.330 | -0.595 |
| Familial Depressive History | -0.627 | -1.125 | -0.386 | -0.622 | -1.780 | -0.398 | -0.618 | -1.520 | 0.358 |
| SES Risk Factors | 2.341 | 0.804 | -0.063 | 2.416 | 1.310 | -0.158 | 2.408 | 1.406 | -0.084 |
| Baseline Depressive Symptoms | NA | | | 0.619 | 0.073 | 0.568 | 0.674 | 0.153 | 0.221 |

*Note.* Depressive symptoms at study enrolment in network 1, at 4-year follow-up in network 2, and at 18-year follow-up in network 3.

**Table S2.**

*Eigenvalues and Proportion of Variance Explained from Exploratory Factor Analysis of Built Environment Indicators.*

| Factor | Eigenvalue | Proportion of Variance | Cumulative Variance |
| --- | --- | --- | --- |
| Factor 1 | 3.939 | 0.492 | 0.492 |
| Factor 2 | 0.982 | 0.123 | 0.615 |
| Factor 3 | 0.916 | 0.115 | 0.730 |
| Factor 4 | 0.672 | 0.084 | 0.814 |
| Factor 5 | 0.659 | 0.082 | 0.896 |
| Factor 6 | 0.379 | 0.047 | 0.943 |
| Factor 7 | 0.319 | 0.040 | 0.983 |
| Factor 8 | 0.134 | 0.017 | 1.000 |

**Table S3.**

*Model fits compared across different factor structure models.*

| Model | CFI | TLI | RMSEA | SRMR |
| --- | --- | --- | --- | --- |
| 1-Factor | 0.825 | 0.755 | 0.179 | 0.057 |
| 2-Factor | 0.933 | 0.834 | 0.224 | 0.038 |
| 3-Factor | 0.927 | 0.86 | 0.155 | 0.051 |

*Note.* CFI = Comparative Fit Indices , TLI = Tucker-Lewis Indices, RMSEA = Root Mean Squared Errors of Approximation, SRMR = Standardised Root Mean Square Residuals.

**Figure S10.**

*Scree plot*

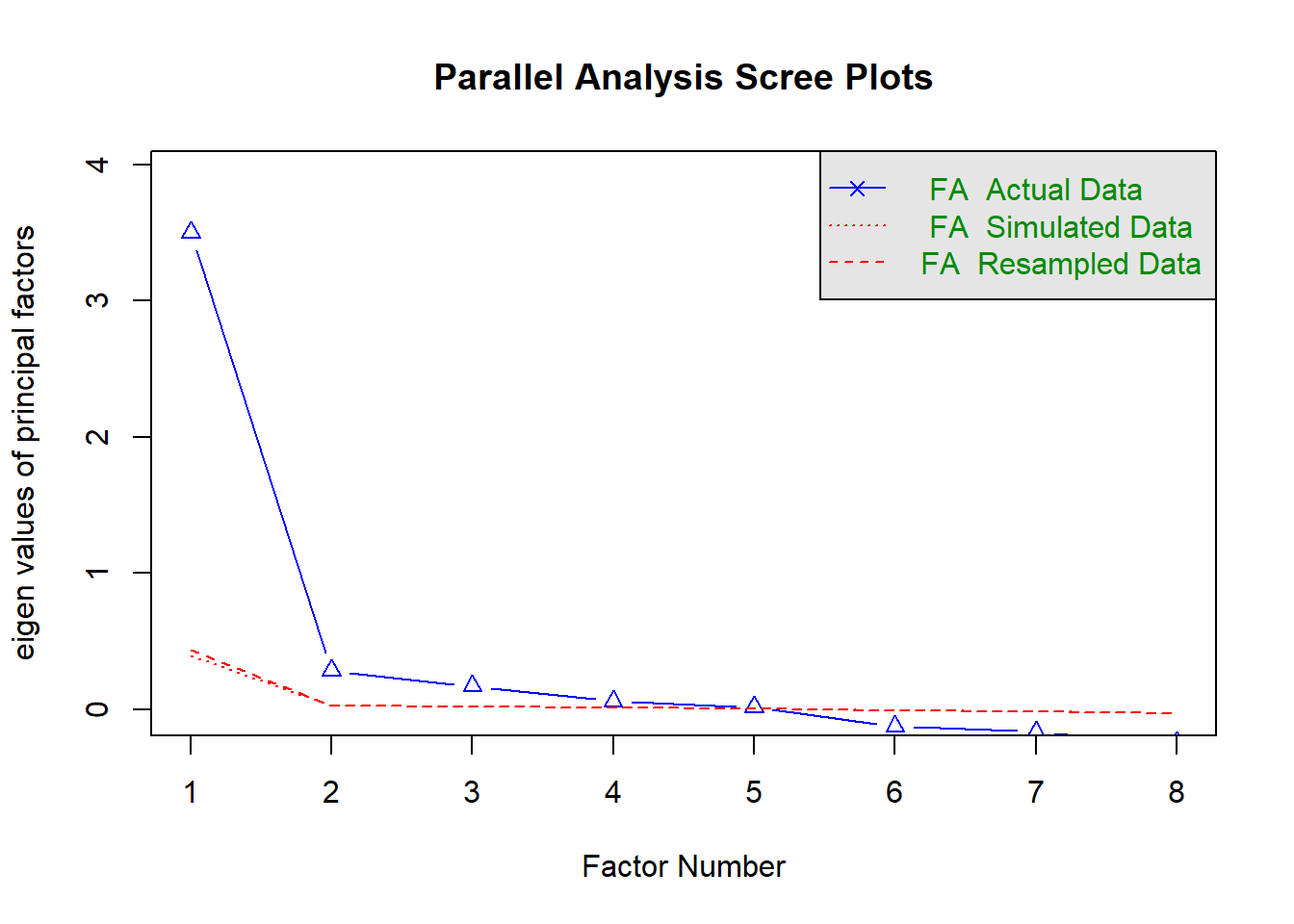

To compare alternative factor structures, we evaluated 1-, 2-, and 3-factor models using multiple fit indices (CFI, TLI, RMSEA, SRMR). As shown in Table S3, all models demonstrated acceptable SRMR values (< .08), whereas CFI, TLI, and RMSEA varied across structures. Although the 2- and 3-factor models showed slightly higher CFI values, the overall pattern of fit indices was comparable across models. To balance parsimony, interpretability, and model fit, we selected the 1-factor model for subsequent regression analyses.

SM 1.1

Built environment measures

*Building Density*

Building density was measured at study enrolment by the number of m^2^ of buildings per km^2^. A 300m buffer score was used, representing the average building density within a 300m radius of the home address. A 300m buffer is commonly used in literature, however there are high correlations amongst different distance buffers and health behaviours ^1–4^. A 300m buffer strikes a balance between capturing a sufficient portion of the built environment with a likelihood of the participant interacting with that portion of their environment.

*Connectivity Density*

Connectivity density at a 300m buffer was measured at study enrolment. This is the number of road intersections, where two or more roads connect in some way, either through meeting or diverging ^5^, per km^2^. Road intersections are places.

*Facility Density*

Facility density at a 300m buffer was measured at study enrolment, through the number of facilities per km^2^, including educational facilities such as schools, health facilities including hospitals, recreational spaces, commercial amenities, and civic services.

*Facility Richness*

Facility richness at study enrolment was measured as the number of different facility types in the 300m buffer (e.g., educational, health) available divided by the maximum potential number of facility types. It was defined through a score of 0 to 1, where greater scores indicate greater facility richness.

*Bus Lines Length*

At study enrollment, the meters of bus lines in length per km^2^ within a 300m radius of participants’ addresses was measured.

*Distance to nearest road*

At study enrolment, the inverse distance in meters to the nearest road from the participants’ home addresses was quantified.

*Walkability*

An average walkability index score (0-1) at study enrolment was measured at a 300m buffer. The score was defined as the mean of deciles for facility richness index, connectivity density, land use diversity and population density ^6,7^. The walkability index containing some components of built environment features included in the analysis, such as facility richness and population density. However, it is recommended by Descarpentrie and colleagues^3^ to use both the walkability scores and the component variables (facility richness index etc.) in order to capture any potential information losses that can occur from transforming data into a single composite score. Greater scores of walkability are strongly associated with increased walking behaviour ^8^.

*Green Space*

Green space at study enrolment was quantified using the Normalized Difference Vegetation Index at a 500m buffer. The 500m buffer was selected as this was the maximum buffer radius available, and greater NDVI buffer sizes have been found to have stronger associations with health than smaller sizes ^9^. Greater buffer sizes are also important as individuals often report travelling substantially further from their home addresses specifically to reach higher quality green spaces, particularly for recreational trips ^10^. Scores range from -1 to +1, with greater scores indicating more green space.

*Population Density*

Population density, the number of inhabitants per km^2^, was measured at study enrolment.

SM 1.2

Covariates

*Contextual Socioeconomic Status (SES) Risk Factors*

SES risk factors scores – comprise of mother’s education, experience of reduced income, losing a job, becoming homeless, and financial difficulties – are measured at study enrolment. Education was coded, in order of highest to lowest SES risk factors, as ‘CSE’, ‘Vocational’, ‘O level’, ‘A level’, and ‘Degree’. Experiences of reduced income, becoming homeless and financial difficulties, in order of highest to lowest SES risk factors, are coded as ‘Affected a lot’, ‘Moderately affected’, ‘Mildly affected’, ‘No effect at all’, and ‘Did not happen’. These are measured at study enrolment.

*Length of Time at Address*

At study enrolment participants reported the number of years since they moved house.

*Familial Depressive History*

Familial depressive history (e.g., known lifetime experience of depressive symptoms or consistent low mood in the participants’ parents) was reported by participants at study enrolment (2=both parents, 1=one parent, 0=none).

*Smoking*

Smoking was self-reported at study enrolment, and responses measured whether participants had never smoked, were a former smoker, or were still currently smoking.

*Age at Depressive Symptoms*

In the same questionnaire participants reported depressive symptoms, they also self-reported age.

*Age at Built environment*

ALSPAC built environment data was collated from multiple external sources and retrospectively added to their database using participant addresses. Hence, similarly to others ^11^, we use mothers’ age at study enrolment to capture the age at built environment as they were measured within a12-month window.

SM 1.3

Missing Data Imputation

We selected mothers who had at least 50% data available for urbanicity features (including building density (300m buffer), connectivity density (300m buffer), facility density (300m buffer), facility richness (300m buffer), bus lines length (300m buffer), bus stops (500m buffer), total traffic load (100m buffer), distance to nearest road, walkability (300m buffer), green space (500m buffer) and population density) and depressive symptoms at study enrolment, leaving up to 10,310 mothers available for analysis. We originally intended to also include bus stops and traffic load as urbanicity features, but this was not possible due to too much missing data (99.5% and 91.5% respectively). The MICE R package ^12^ was used to impute missing values, through Random Forest, in built environment (at study enrolment) and depressive symptoms (only at follow-ups) variables, as well as covariates (including maternal age at environment measurement, age at depressive symptoms, familial depressive history, length of time at address, smoking, SES risk factors). Participants were removed if they had more than 50% missingness in built environment variables and no depressive symptom data at study enrolment. Urbanicity features, depressive symptoms at all time points and all covariates were used as predictors in a predictor matrix to impute missing data. Covariates included age at urbanicity measurement, age at depressive symptom measurement, length of time at address, familial depressive history, contextual SES risk and smoking. Auxiliary variables (i.e., variables selected to inform imputation but not used in the analysis) included mothers’ travel behaviour at 2-year follow-up (and age at measurement), child ethnicity (at age 17), mothers’ physical activity (at study enrolment), child sex , child travel behaviour at age 13 (and age at measurement), child physical health (at age 13), child pro-environmentalism (at age 13) and child cognition at age 24 (and age at measurement). Auxiliary variables are variables which are going to be used in later analyses, have been identified as possible moderators or are closely associated with variables in this study in order to reduce bias ^13^.

Imputation was performed using 60 iterations and 30 imputed datasets. The predictor matrix was specified using quickpred(), with a minimum correlation set to 0.05, striking a balance between having sufficient variable breadth yet avoiding weak predictors.

**Table S4.**

*Original data compared to pooled (average across MICE datasets) imputed data (30 imputations).*

|  | **N** | | **Mean** | | **SD** | | **Min** | | **Max** | | **Difference Statistic** | ***P* Value** |
| --- | --- | --- | --- | --- | --- | --- | --- | --- | --- | --- | --- | --- |
|  | Original | Imputed | Original | Imputed | Original | Imputed | Original | Imputed | Original | Imputed |  |  |
| Building Density | 10310 | 10310 | 400733.27 | 400733.27 | 135306.01 | 135306.01 | 20000.00 | 20000.00 | 670000.00 | 670000.00 | t = 0.00 | 0.99 |
| Bus Lines | 9500 | 10310 | 3200.21 | 3143.13 | 1599.47 | 1579.45 | 1000.00 | 1000.00 | 8000.00 | 8000.00 | t = 2.53 | 0.01 |
| Connectivity Density | 10310 | 10310 | 129.73 | 129.73 | 60.19 | 60.19 | 10.00 | 10.00 | 340.00 | 340.00 | t = 0.00 | 0.99 |
| Facility density | 10309 | 10310 | 33.10 | 33.10 | 42.58 | 42.58 | 0.00 | 0.00 | 200.00 | 200.00 | t = -0.01 | 0.99 |
| Facility Richness | 10310 | 10310 | 0.07 | 0.07 | 0.07 | 0.07 | 0.00 | 0.00 | 0.30 | 0.30 | t = 0.00 | 0.99 |
| Green Space | 10310 | 10310 | 0.42 | 0.42 | 0.09 | 0.09 | 0.20 | 0.20 | 0.70 | 0.70 | t = 0.00 | 0.99 |
| Nearest Road | 10310 | 10310 | 0.07 | 0.07 | 0.03 | 0.03 | 0.01 | 0.01 | 0.20 | 0.20 | t = 0.00 | 0.99 |
| Population density | 10169 | 10310 | 4915.82 | 4870.93 | 2740.44 | 2752.60 | 1000.00 | 1000.00 | 10000.00 | 10000.00 | t = 1.13 | 0.26 |
| Walkability | 10072 | 10310 | 0.30 | 0.30 | 0.06 | 0.06 | 0.15 | 0.15 | 0.50 | 0.50 | t = 1.12 | 0.27 |
| Familial history of depression | | | | | | | | | | | | |
| N | 10310 | 10310 | NA | | | | | | | | χ² = 0.00 | 0.99 |
| Both parents | 2.09% | 2.09% |  |  |  |  |  |  |  |  |  |  |
| One parent | 21.41% | 21.41% |  |  |  |  |  |  |  |  |  |  |
| Neither parents | 76.51% | 76.51% |  |  |  |  |  |  |  |  |  |  |
| Ethnicity | | | | | | | | | | | | |
| N | 9500 | 10310 | NA | | | | | | | | χ² = 0.36 | 0.55 |
| White | 97.62% | 97.77% |  |  |  |  |  |  |  |  |  |  |
| Non-White | 2.38% | 2.23% |  |  |  |  |  |  |  |  |  |  |
| Smoking (study enrolment) | | | | | | | | | | | | |
| N | 9063 | 10310 | NA | | | | | | | | χ² = 4.71 | 0.09 |
| Smoking | 19.17% | 19.62% |  |  |  |  |  |  |  |  |  |  |
| Former smoker | 30.21% | 31.27% |  |  |  |  |  |  |  |  |  |  |
| Never smoked | 50.62% | 49.11% |  |  |  |  |  |  |  |  |  |  |
| Length of time at address | 9446 | 10310 | 3.03 | 3.01 | 3.64 | 3.66 | 0.00 | 0.00 | 34.00 | 34.23 | t = 0.43 | 0.67 |
| Contextual SES | 8390 | 10310 | 1.66 | 1.69 | 0.47 | 0.48 | 1.00 | 1.00 | 4.25 | 4.25 | t = -4.60 | < .001 |
| Age at birth | 10310 | 10310 | 28.11 | 28.11 | 4.83 | 4.83 | 15.00 | 15.00 | 44.00 | 44.00 | t = 0.00 | 0.99 |
| Age at depressive symptoms (study enrolment) | 10292 | 10310 | 27.72 | 27.73 | 4.83 | 4.84 | 13.00 | 13.00 | 44.00 | 44.00 | t = -0.02 | 0.99 |
| Age at depressive symptoms (4-year follow-up) | 7485 | 10310 | 32.01 | 31.53 | 4.61 | 4.74 | 19.00 | 18.67 | 52.00 | 53.73 | t = 6.83 | < .001 |
| Age at depressive symptoms (18-year follow-up) | 3324 | 10310 | 48.52 | 47.26 | 4.38 | 4.60 | 34.00 | 34.00 | 63.00 | 63.00 | t = 14.26 | < .001 |
| Depressive Symptoms (study enrolment) | 10310 | 10310 | 4.46 | 4.46 | 3.09 | 3.09 | 0.00 | 0.00 | 16.00 | 16.00 | t = 0.00 | 0.99 |
| Depressive symptoms (4-year follow-up) | 7489 | 10310 | 4.17 | 4.24 | 3.21 | 3.24 | 0.00 | 0.00 | 16.00 | 16.00 | t = -1.39 | 0.17 |
| Depressive Symptoms (18-year follow-up) | 2318 | 10310 | 14.70 | 14.66 | 4.36 | 4.36 | 3.00 | 3.00 | 25.00 | 25.00 | t = 1.17 | 0.24 |

SM 1.4

Network analysis methods

In our pre-registration we specified a moderate EBIC tuning parameter of 0.5. However, during data analysis a value of 0.4 was ultimately selected as it struck a better balance between specificity and conservativity ^14^. Edges first get selected or removed by LASSO, and then the EBIC tuning parameter shrinks weaker edges towards zero. Greater EBIC tuning parameters shrink towards zero more aggressively. A parameter of 0.4 excluded weak edges to reduce noise, whilst still retaining sufficient sensitivity to detect important edges ^15^.

Next, we conducted a multiple regression testing whether the single built environment factor predicted depressive symptoms at study enrolment, whilst controlling for age at built environment measurement, age at depressive symptom measurement, length of time at address, familial depressive history, contextual SES risk and smoking. Then we tested whether the built environment factor predicted depressive symptoms at a 4-year follow up, whilst controlling for age at built environment measurement, age at depressive symptom measurement, length of time at address, familial depressive history, contextual SES risk, smoking and baseline depressive symptoms. Lastly, we tested whether a built environment factor predicted depressive symptoms at an 18-year follow-up, whilst controlling for age at built environment measurement, age at depressive symptom measurement, length of time at address, familial depressive history, contextual SES risk, smoking and baseline depressive symptoms.
